# A survey of experts to identify methods to detect problematic studies: Stage 1 of the INSPECT-SR Project

**DOI:** 10.1101/2024.03.18.24304479

**Authors:** Jack Wilkinson, Calvin Heal, George A Antoniou, Ella Flemyng, Alison Avenell, Virginia Barbour, Esmee M Bordewijk, Nicholas J L Brown, Mike Clarke, Jo Dumville, Steph Grohmann, Lyle C. Gurrin, Jill A Hayden, Kylie E Hunter, Emily Lam, Toby Lasserson, Tianjing Li, Sarah Lensen, Jianping Liu, Andreas Lundh, Gideon Meyerowitz-Katz, Ben W Mol, Neil E O’Connell, Lisa Parker, Barbara Redman, Anna Lene Seidler, Kyle Sheldrick, Emma Sydenham, Darren L Dahly, Madelon van Wely, Lisa Bero, Jamie J Kirkham

## Abstract

**Background:** Randomised controlled trials (RCTs) inform healthcare decisions. Unfortunately, some published RCTs contain false data, and some appear to have been entirely fabricated. Systematic reviews are performed to identify and synthesise all RCTs which have been conducted on a given topic. This means that any of these ‘problematic studies’ are likely to be included, but there are no agreed methods for identifying them. The INSPECT-SR project is developing a tool to identify problematic RCTs in systematic reviews of healthcare-related interventions. The tool will guide the user through a series of ‘checks’ to determine a study’s authenticity. The first objective in the development process is to assemble a comprehensive list of checks to consider for inclusion.

**Methods:** We assembled an initial list of checks for assessing the authenticity of research studies, with no restriction to RCTs, and categorised these into five domains: Inspecting results in the paper; Inspecting the research team; Inspecting conduct, governance, and transparency; Inspecting text and publication details; Inspecting the individual participant data. We implemented this list as an online survey, and invited people with expertise and experience of assessing potentially problematic studies to participate through professional networks and online forums. Participants were invited to provide feedback on the checks on the list, and were asked to describe any additional checks they knew of, which were not featured in the list.

**Results:** Extensive feedback on an initial list of 102 checks was provided by 71 participants based in 16 countries across five continents. Fourteen new checks were proposed across the five domains, and suggestions were made to reword checks on the initial list. An updated list of checks was constructed, comprising 116 checks. Many participants expressed a lack of familiarity with statistical checks, and emphasized the importance of feasibility of the tool.

**Conclusions:** A comprehensive list of trustworthiness checks has been produced. The checks will be evaluated to determine which should be included in the INSPECT-SR tool.

## Background

Randomised controlled trials (RCTs) are performed to investigate whether treatments are safe and effective. Systematic reviews exploring health interventions aim to include all relevant RCTs, appraising and synthesising this evidence to arrive at an overall conclusion about whether an intervention works and whether it causes harm. Problematic studies pose a threat to the evidence synthesis paradigm. These are defined by Cochrane as “any published or unpublished study where there are serious questions about the trustworthiness of the data or findings, regardless of whether the study has been formally retracted”(1, 2). Studies may be problematic because they include some false data or results, or may be entirely fabricated. Research misconduct is just one possible explanation for false data. Another possibility would be the presence of catastrophic failures in the conduct of the study, such as miscoding of patient conditions (e.g., inverting active treatment and placebo conditions), failure in the computerised randomisation service, or severe errors in the analysis code. Whether they are the result of deliberate malpractice or honest error, these issues may not be immediately apparent to journal editors and peer reviewers. Consequently, problematic studies may be published, and subsequently included in systematic reviews. Studies are routinely appraised on the basis of their methodological validity during the systematic review process. However, these assessments are predicated on the assumption that the studies and the data they are based on are authentic, and also that the authors did not make any major errors during data collection, analysis or reporting. In fact, many reports of problematic studies describe sound methodology, and so are not flagged by critical appraisal tools. At present, there are no agreed methods for identifying problematic RCTs, and it is typical for no assessment of authenticity to be undertaken at all. This means that there are no processes for preventing problematic RCTs from being included in systematic reviews, distorting the clinical evidence base, and potentially leading to harm.

This prompts the question of how we can systematically detect problematic studies. The overall aim of the INSPECT-SR (INveStigating ProblEmatic Clinical Trials in Systematic Reviews) project is to develop and evaluate a tool for identifying problematic studies in the context of systematic reviews of RCTs of health interventions(3). The INSPECT-SR tool will guide the user through a series of ‘checks’ for study trustworthiness. The development approach involves identifying a comprehensive list of checks for trustworthiness, and subjecting these to evaluation to determine which to include in the tool. The first objective in this process is generation of a comprehensive list of possible trustworthiness checks for evaluation in subsequent stages of the project. In addition to its use in the development of INSPECT-SR, we anticipate that this comprehensive list of trustworthiness checks will be a useful contribution to the research integrity literature.

The aim of Stage 1 of the INSPECT-SR process, reported here, was to assemble a comprehensive list of checks for potentially problematic studies, using a survey of experts and people with relevant experience. Specific objectives were to identify hitherto unidentified checks and to obtain feedback on previously identified ones.

## Methods

The methods used in this study have been described in an online protocol (https://osf.io/6pmx5/) and in a protocol paper describing the INSPECT-SR project (3). We give an overview here.

### Assembling an initial list of checks for problematic studies

We assembled an initial list of trustworthiness checks of research studies, using several sources. Although our long-term goals in the INSPECT-SR project are to develop a tool for assessing RCTs in particular, at this stage we did not restrict the list to checks which had been proposed specifically in an RCT context. This was to ensure that we did not miss checks which could potentially be of use for assessing RCTs. However, some checks were considered as being out of scope (e.g. they referred to purchasing of animals in animal studies, or related to risk of bias (4)). Excluded checks are shown in the Supplementary Material. We included checks which appeared in a recent scoping review (5) and qualitative study of experts (6). We located and read the original studies or reports described by the scoping review to ensure that no checks were omitted. For example, the scoping review included the REAPPRAISED checklist (7) and we extracted the individual items from that checklist and included them in our list. We added additional checks which were known to the research team. For example, JW has a background in undertaking integrity investigations for journals and publishers, and he added checks used in this work. We started by including the checks from the papers included in the scoping review before adding any additional checks included in the qualitative study, and finally any additional checks known to the author team. If the same check was encountered multiple times during this process, it was added to the list only once. Some checks were considered redundant given other checks, and were excluded on this basis (see excluded checks in Supplementary Material, (5–10)). We defined five preliminary domains and categorized each check into one of these domains. The domains used were Inspecting results in the paper, Inspecting the research team, Inspecting conduct, governance and transparency, Inspecting text and publication details, and Inspecting individual participant data. The wording and categorization of the checks was reviewed by the project Expert Panel (3) and revised accordingly. The majority were rephrased as questions for consistency.

### Online survey

The initial list of checks was implemented as an online survey in Qualtrics (11). The survey can be viewed at https://osf.io/s34hx. Participants were informed about the motivation for the study and the content of the survey should they choose to participate. The survey then asked participants about their experience in assessing potentially problematic studies (with these questions being used to confirm eligibility), and presented participants with the list of checks that could be used to assess potentially problematic studies. The checks were presented in their preliminary domains, and both the order of domains and the order of checks within each domain were randomised, to minimise the impact of potential sequence effects. Each check was presented alongside a free-text box, and participants were advised to comment on any aspect if they wished to do so. At the end of the list, participants were asked whether they were aware of any other checks which had not featured on the list, and were presented with a free text box to describe these. The survey was piloted by members of the research team and colleagues prior to launch. The survey opened on 14th November 2022 and closed on 25th January 2023. The survey was anonymous – we did not collect any identifying information in the survey. Ethical approval was not required for this study, since it involved asking experts for their professional opinion.

### Participants

People with expertise or experience of assessing potentially problematic studies, either prior to or post-publication, were eligible to participate in the survey. This included editors of health journals, research integrity professionals, and researchers with experience of conducting research integrity investigations, or of undertaking related methodological research.

We implemented a multifaceted recruitment strategy. We promoted the project via conferences (International Clinical Trials Methodology Conference 2022, International Congress on Peer review and Scientific Publication 2022), social media (Twitter account of JW), and via a group of researchers and publishing representatives established to discuss problems posed by paper mills (12), inviting potential participants to contact JW. We identified and contacted individuals involved in relevant research integrity activities, including researchers, journal editors, and research integrity professionals. Additionally, the INSPECT-SR working group includes a Steering Group and an Expert Advisory Panel (3), and members of both of these were invited to participate if they met the eligibility criteria (the authors of the present article represent members of both groups). We invited eligible individuals by personalised email, and asked whether they could suggest any other potential participants. We aimed for a geographically diverse sample, and monitored responses to the question ‘In which country do you primarily work?’ as responses accrued. We made efforts to identify and invite potential participants based in nations which were not represented by reaching out to professional contacts in those regions and asking for suggestions for potential participants, and also by asking for suggestions from the organizers of recent and upcoming World Conferences on Research Integrity. We also identified international research integrity networks and contacted them to request details of the project to be shared with their members (African Research Integrity Network, Association for the Promotion of Research Integrity), again with a request for potential participants to contact JW.

### Sample size

We targeted a minimum sample size of 50 participants, and did not end recruitment once this target was met, first because our goal was to obtain feedback from as many experts as possible within the available timeframe, and second because we did not perform any inferential statistical analyses. The sample size was largely based on pragmatic considerations – we believed 50 participants was realistic based on previous research in similar populations e.g. (13) while representing a sufficient number of responses to obtain thorough feedback on the list of the checks.

### Statistical analysis

We examined survey results, including participant characteristics, using descriptive statistics. Additional items suggested by respondents, and comments made on existing items, were summarised. The survey responses were used to add further items to the list, and to amend the wording of existing items, subject to review by Steering Group and Expert Advisory panel members.

## Results

The initial list entered into the survey contained 102 checks (76 from papers referenced by the scoping review, 14 from the qualitative study, and 12 additional checks suggested by the author team). Figure 1 shows the distribution of the checks across the five domains. Eighty individuals accessed the survey. Nine individuals did not meet the eligibility criteria (insufficient experience in assessing problematic studies). Consequently, responses were obtained from 71 participants. The study dataset is available at https://osf.io/6pmx5/.

**Figure 1:**
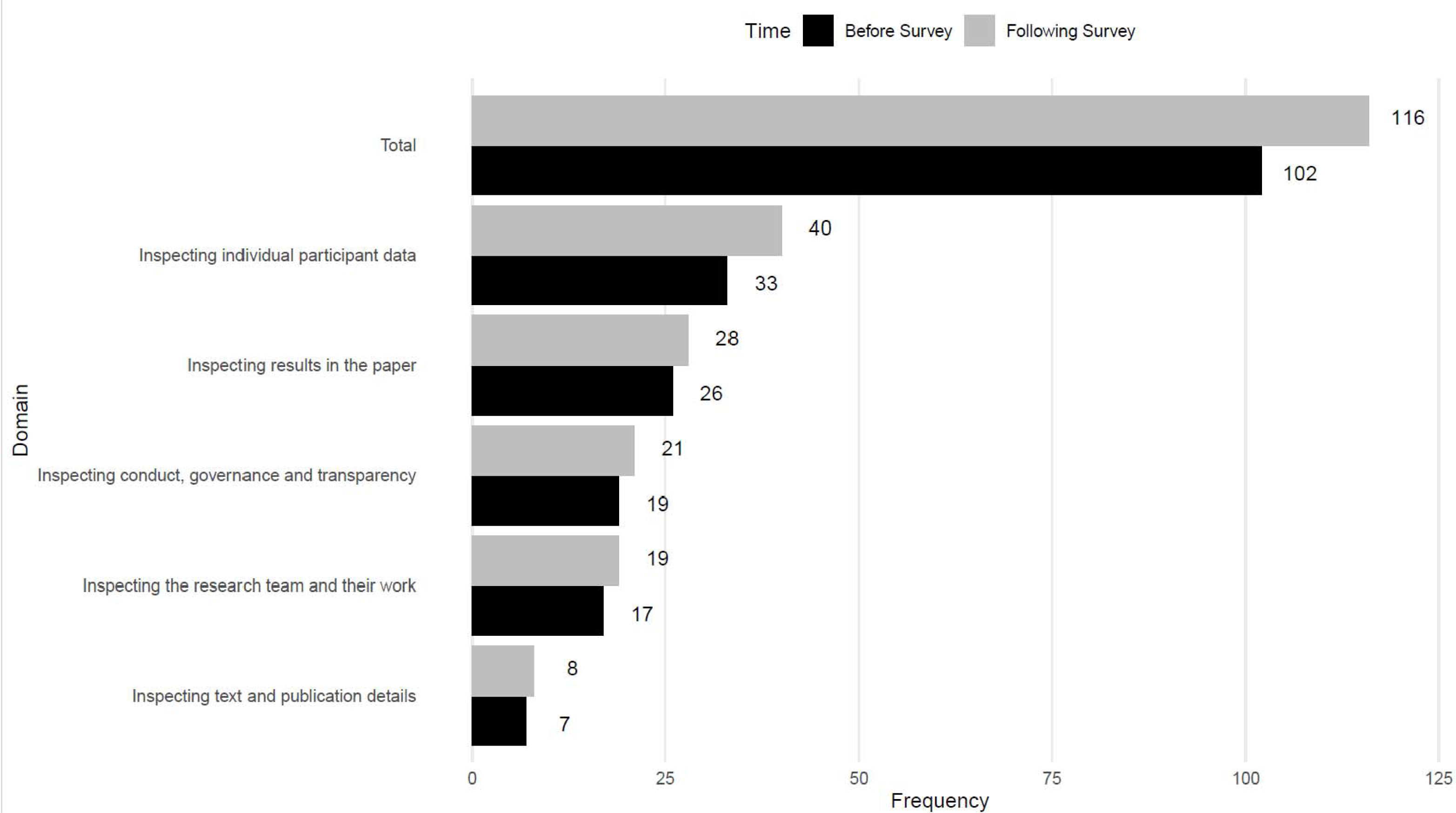
Number of checks in each domain before and after the survey.

### Characteristics of participants

Table 1 shows the characteristics of participants. Responses were obtained from participants based in 16 countries across five continents, although the majority (55%) of participants were based in Europe (Table 1). The experience of the included participants is also outlined in Table 1. The majority had assessed potentially problematic studies as an independent researcher (85%) with around half having done so as a peer reviewer (49%). Most had been involved in methodological research into identifying problematic studies (58%), noting that this could have referred to involvement in the INSPECT-SR project. Fewer participants had investigated potentially problematic studies as a journal editor (28%) or research integrity professional (27%).

**Table 1:** Characteristics of participants. Frequency (%) *Multiple responses permitted

| Characteristic | N (%) |
| --- | --- |
| Primary location of work |  |
| Europe | 39 (55%) |
| Australia/Oceania | 15 (21%) |
| North America | 10 (14%) |
| Africa | 5 (7%) |
| South America | 1 (1%) |
| Missing | 1 (1%) |
| Experience* |  |
| Have you assessed potentially problematic studies as an independent researcher (post-publication)? | 60 (85%) |
| Have you conducted methodological research into the issue of identifying problematic studies? | 41 (58%) |
| Have you assessed potentially problematic studies as a peer reviewer (pre-publication)? | 35 (49%) |
| Have you assessed potentially problematic studies as a journal editor? | 20 (28%) |
| Have you assessed potentially problematic studies in any other capacity not listed here? | 20 (28%) |
| Have you assessed potentially problematic studies as a research integrity professional? | 19 (27%) |
| Have you assessed potentially problematic studies at the request of a journal or publisher? | 17 (24%) |
| Have you assessed potentially problematic studies you have been involved in (e.g. possible misconduct by collaborators)? | 10 (14%) |

### Feedback on existing checks

The full list of comments by item on the list can be found in the Supplementary Material. Many suggestions revolved around specific wording changes to checks to clarify their purpose and differentiate them from each other. Feedback indicated that some checks were not well understood by participants. As an example, one check included in the domain Inspecting individual participant data was to ‘make star plots for each group’(10, 14). This check received eight separate comments detailing participants’ unfamiliarity with this concept. Similar comments were made in relation to many of the statistical checks included on the list, both in the aforementioned domain and also in the domain Inspecting results in the paper. Some comments indicated that the domain name Inspecting the research team did not clearly correspond to some of the checks contained in the domain, which referred to checking other work conducted by the research team of the index study.

### Proposal of new checks

There were 38 suggestions of checks to add to the list. We were unable to interpret the meaning of four suggestions. Of the remainder, 19 suggestions, describing 14 distinct checks, were considered novel, that is, not sufficiently similar to existing checks to be considered a duplication. (Table 2, with wordings edited for clarity). We categorized the proposed checks. We considered seven (50%) of the novel checks to fall within the Inspecting individual participant data domain. It was proposed that the country in which the study was conducted be included as a check. We have included this in Table 2 for completeness, and discuss the implications of this check in the discussion.

**Table 2.**
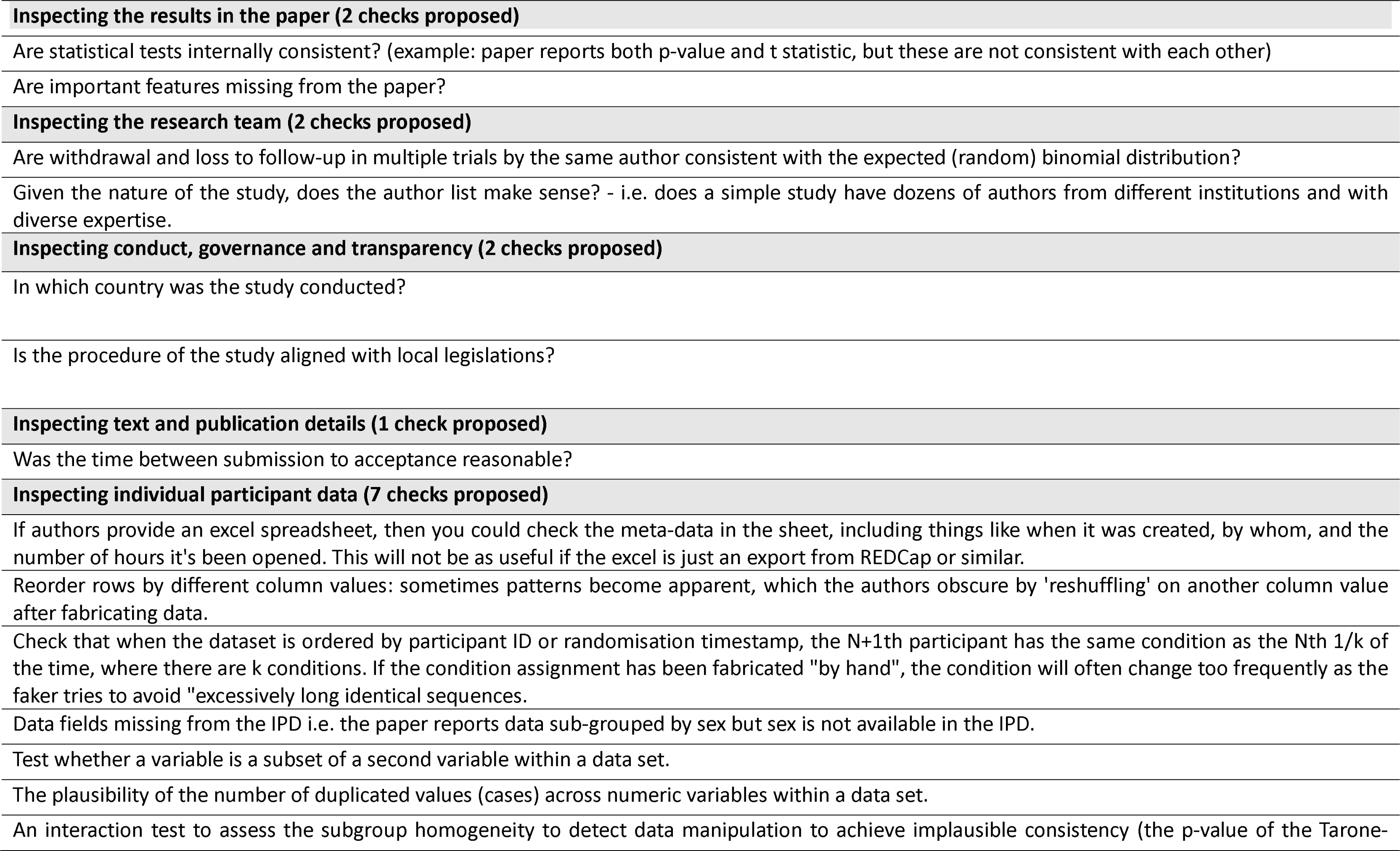
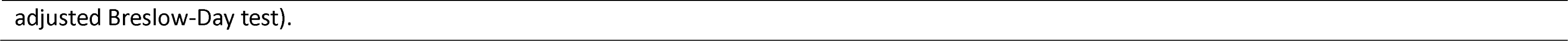
Novel suggestions for checks for problematic studies.

### General feedback

Finally, participants were offered the chance to comment on the survey, or on the topic more generally. Redacted versions of these comments are included in the Supplementary Material. Redaction has been performed to conceal the identities of the participants and of the subjects of their comments. Desire for a practical, short tool was a common theme, with several participants suggesting it should be structured so that easier checks are performed first. If the outcome of these checks proved definitive (e.g. identifying or assuaging serious concerns), this would avoid the use of more burdensome or complex methods appearing later in the tool.

### Updated list of checks

Based on the responses to the survey, an updated list of possible checks for potentially problematic studies was developed, incorporating the new suggestions and updating the wording of items in response to feedback. The number of items following the survey is shown in Figure 1, and the updated list is shown in the Supplementary Material (7, 9, 10, 14-42). Figure 2 shows the origin of checks included in the final list. In response to survey feedback, we changed the second domain name to Inspecting the research team and their work.

**Figure 2:**
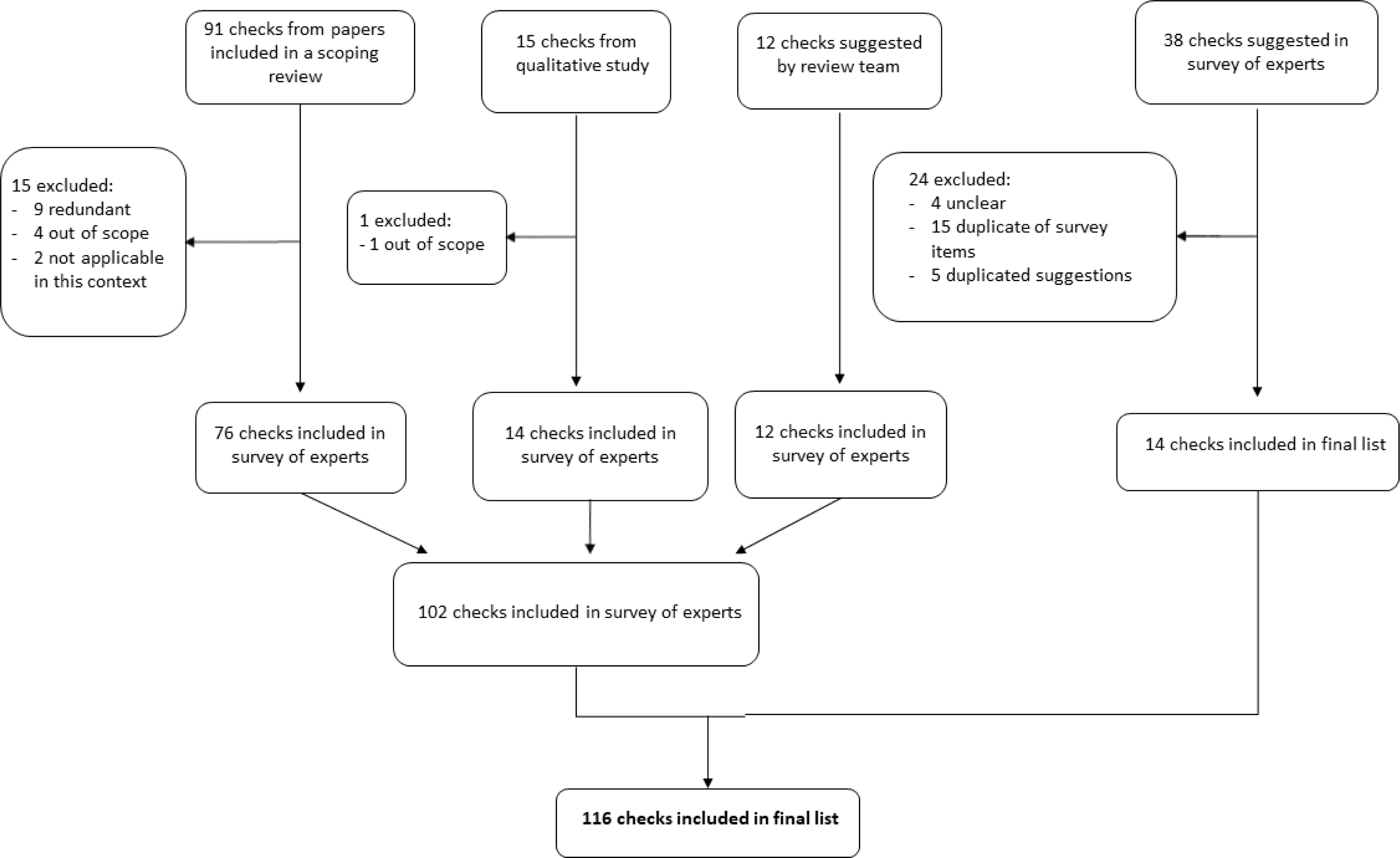
Flow chart showing origin of checks included in final list.

## Discussion

We conducted an international survey of experts to elaborate an extensive list of potential checks for identifying problematic studies. The items on the list will be evaluated for their usefulness and feasibility to determine which checks should be included in the INSPECT-SR tool and any implications for the tool’s structure (3). It should be emphasised that a check’s inclusion on the list does not amount to an endorsement by the research team. We anticipate that many of these checks will ultimately be found to be infeasible or simply not informative.

Participant responses highlighted a number of important considerations for the development of a tool for assessing potentially problematic studies. Despite representing a cohort of individuals with experience and expertise in problematic studies, many respondents expressed a lack of familiarity with items included on the list, particularly those relating to statistical methods. Given that the INSPECT-SR tool is intended for use by researchers without this level of expertise, our findings suggest that these checks would need to be accompanied by clear guidance to facilitate use and prevent misapplication and misinterpretation, similar to explanation and elaboration documents created to accompany reporting guidelines (43, 44), or that application of these checks might need input from a statistician. This may also need to be accompanied by software to facilitate the implementation of more complex checks. In addition, this suggests that clear explanations would be needed to allow the checks to be evaluated as part of a subsequently planned consensus process (3). Another clear theme among the survey responses related to the need for a tool to be feasible in terms of the time required to implement it. Some respondents expressed concern about the prospect of a tool involving too many checks; some had mistaken the list to represent the proposed tool, noting that it would not be workable. These concerns highlight the importance of evaluating not only the feasibility of individual items but also the practicality of the resulting tool. To this end, a draft version of the tool will be extensively tested in the production of new systematic reviews of RCTs, and revised accordingly. One proposal to increase the viability of the tool was to arrange the checks in a hierarchical format, with initial, less burdensome checks being performed first, potentially obviating more difficult checks should clear problems be apparent.

We included some checks which can only be applied when the underlying individual participant data are available in the survey. Often, these data will not be available to researchers, and so these checks will not be possible. This suggests that the core INSPECT-SR tool should not include checks requiring individual participant data. Accordingly, we will develop an extension to the core tool (working title INSPECT-IPD) which may be applied when the underlying dataset is available. Checks in the individual participant data domain were also unfamiliar to many participants, suggesting that the development of this extension would require input from subspecialists in forensic statistics.

One check which was proposed in response to the survey was to consider the country in which the study was performed. The introduction of this check would be contentious. From an empirical standpoint, while it is plausible that research misconduct would be more likely to occur in settings with limited research governance and oversight, robust evidence relating to the geographical variation in prevalence of problematic studies is relatively limited (with some exceptions, e.g. (45, 46)). From an ethical standpoint, using the country of origin as an indicator of study provenance in its own right would discriminate against honest researchers based in these locations. This check will be subjected to evaluation as part of the development process.

A considerable limitation of the present study is the failure to recruit many participants situated outside of Europe, Australia, and North America. Improving geographical representation in subsequent stages of the project will be necessary to ensure that the tool is both equitable and useful for the assessment of research globally. Some responses described concerns that some checks could not be reliably performed without knowledge of the local context. We also acknowledge that it is possible some checks have not been identified, and so we will ask participants in a subsequent Delphi exercise to propose any additional suggestions for evaluation to minimize the likelihood anything important is missed.

The items on the list will be evaluated via an application of the items on the list to RCTs in 50 Cochrane Systematic Reviews, an online Delphi survey, and consensus meetings, to produce a draft version of the INSPECT-SR tool. The draft version will then be subject to testing by users, and feedback from this testing will be used to improve and finalize the tool (3). The final version will represent a feasible tool, backed by empirical evidence and broad expert consensus, for evaluating potentially problematic studies in health-related systematic reviews.

## Data Availability

The study dataset is available at https://osf.io/6pmx5/

## Ethical approval

The University of Manchester ethics decision tool was used on 30/09/22. Ethical approval was not required for this study, since it involved asking experts for their professional opinion.

## Funding

This study/project is funded by the NIHR Research for Patient Benefit programme (NIHR203568). The views expressed are those of the author(s) and not necessarily those of the NIHR or the Department of Health and Social Care.

## Declaration of interests

JW, CH, GAA, LB, JJK declare funding from NIHR (NIHR203568) in relation to the current project. JW additionally declares Stats or Methodological Editor roles for BJOG, Fertility and Sterility, Reproduction and Fertility, Journal of Hypertension, and for Cochrane Gynaecology and Fertility. CH declares a Statistical Editor role for Cochrane Colorectal. LB additionally declares a role as Academic Meta-Research Editor for PLoS Biology, and that The University of Colorado receives remuneration for service as Senior Research Integrity Editor, Cochrane. JJK additionally declares a Statistical Editor role for The BMJ. AA declares that The Health Services Research Unit, University of Aberdeen, is funded by the Health and Social Care Directorates of the Scottish Government. VB is EiC of the Medical Journal of Australia and on the Editorial Board of Research Integrity and Peer Review. NJLB declares roles as Editorial Board member for International Review of Social Pyschology/ Revue Internationale de Psychologie Sociale, Statistical Advisory Board member for Mental Health Science, and Advisory Board member for Meta-Psychology. MC declares that he is Co-ordinating Editor for the Cochrane Methodology Review Group, Editor in Chief, Journal of Evidence-Based Medicine, and Coordinating Editor, James Lind Library. EF, SG and TLa declare employment by Cochrane. EF additionally declares a role as Editorial Board member for Cochrane Synthesis and Methods. TLa additionally declares authorship of a chapter in the Cochrane Handbook for Systematic Reviews of Interventions and that he is a developer of standards for Cochrane intervention reviews (MECIR). TLi is funded by the National Eye Institute, National Institutes of Health (Grant #UG1 EY020522). SL is funded by NHMRC (APP1195189), and holds general or methodological editor positions for Cochrane Gynaecology and Fertility, Fertility and Sterility, and Human Reproduction. AL is on the editorial board of BMC Medical Ethics. BWM declares roles as Editor for Cochrane Gynaecology and Fertility and Sexually Transmitted Infections and for Fertility and Sterility. SL declares roles as Associate Editor for Human Reproduction, Methodological Editor for Fertility and Sterility, and Editor for Cochrane Gynaecology and Fertility. NOC is a member of the Cochrane Editorial Board and holds an ERA-NET Neuron Co-Fund grant for a separate project. ALS declares funding from Australian National Health and Medical Research Council Investigator Grants (GNT2009432). ES is a Sign-off Editor for the Cochrane Library. MvW is coordinating editor of Cochrane Gynaecology and Fertility and Cochrane Sexually Transmitted Infections, Methodological Editor of Human Reproduction Update and editorial Editor of Fertility and Sterility. All other authors have nothing to declare.

## Acknowledgements

The authors would like to thank Richard Stevens for helpful comments during the planning of this study.

## SUPPLEMENTARY MATERIAL

**Section A. List of comments.**

**Section B. Updated list of checks for problematic studies**

**Section C. Items excluded from the survey.**

### Section A. List of comments. Note that identifying information has been redacted

#### Inspecting the results in the paper

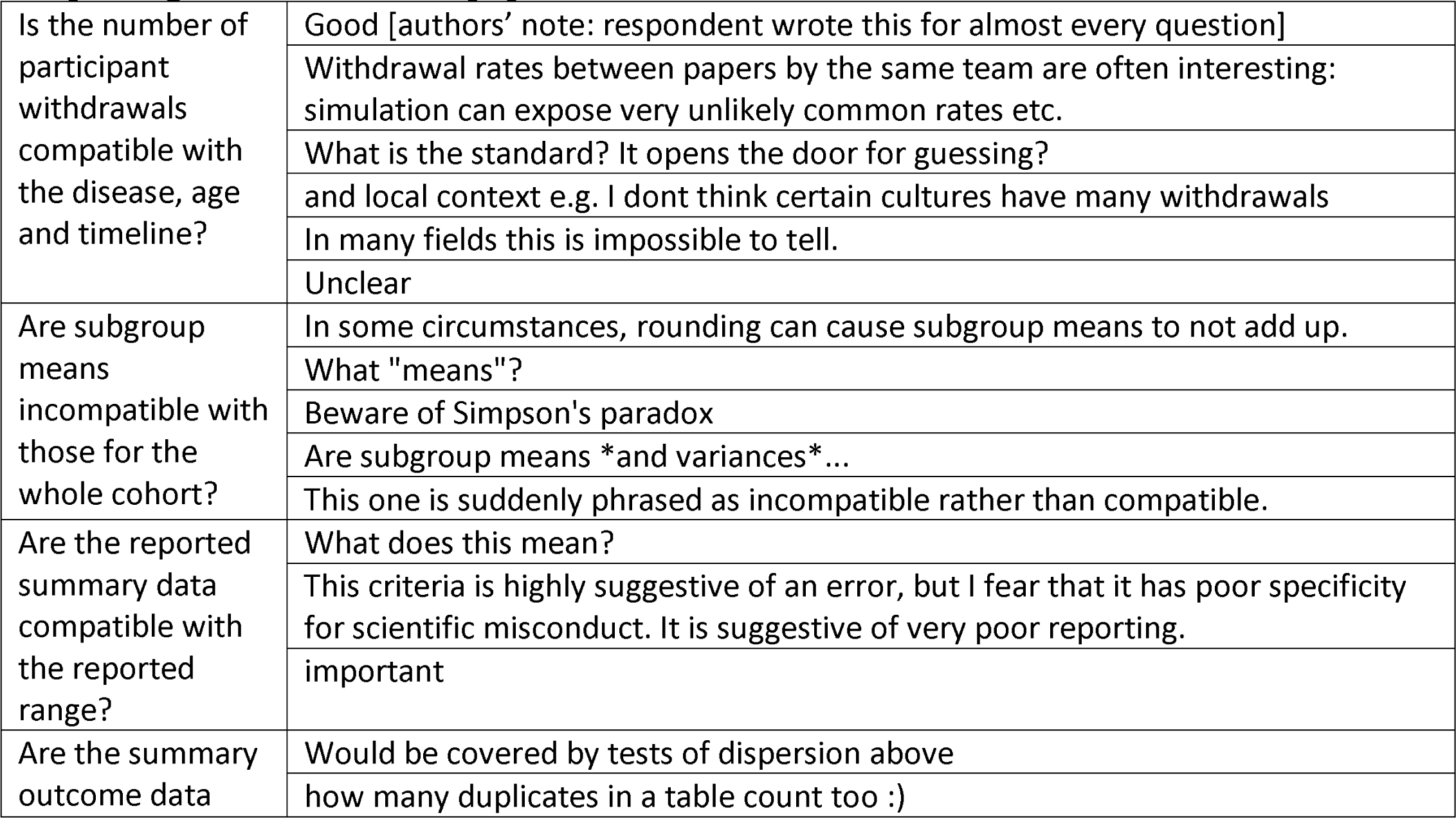
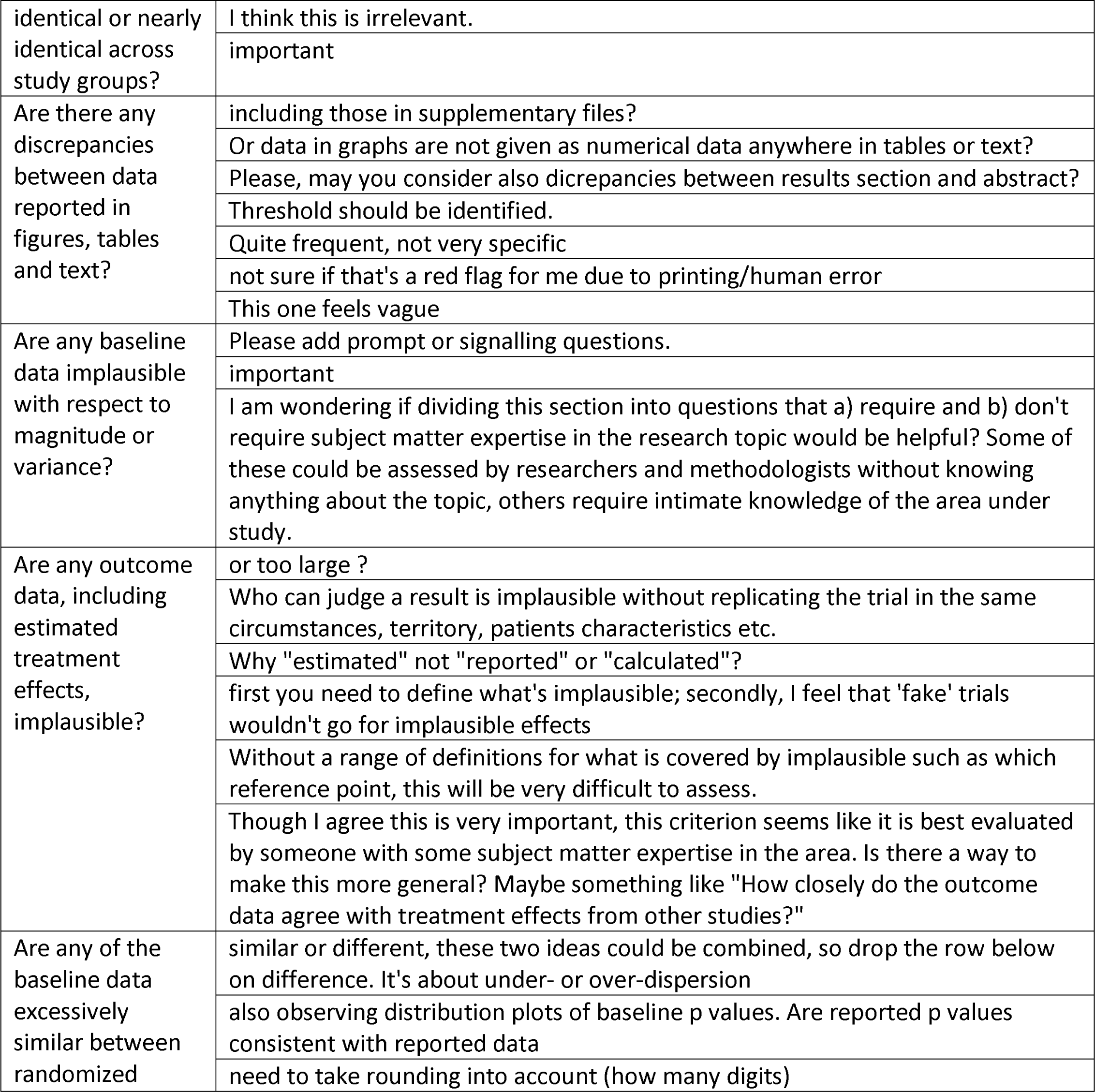
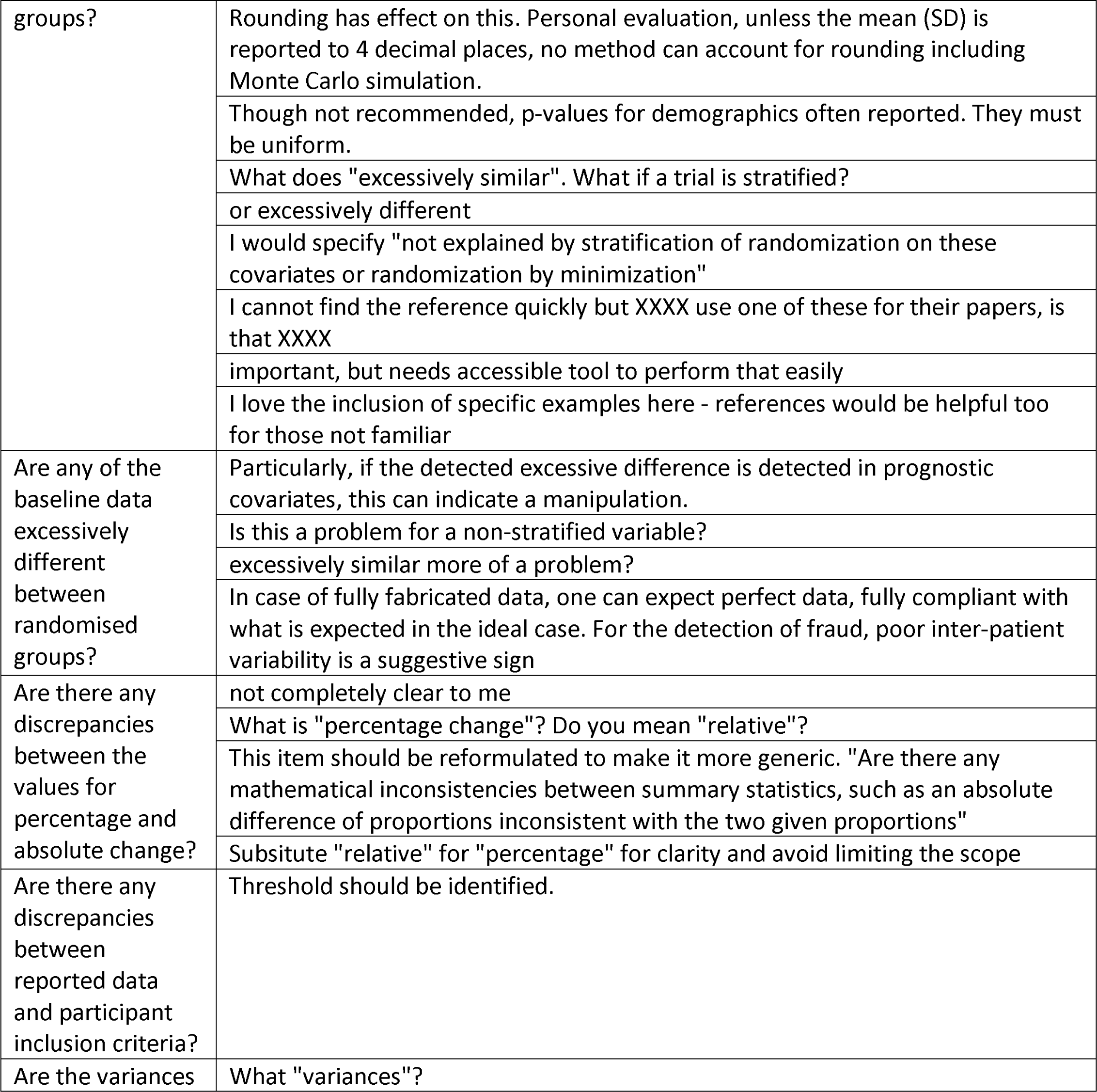
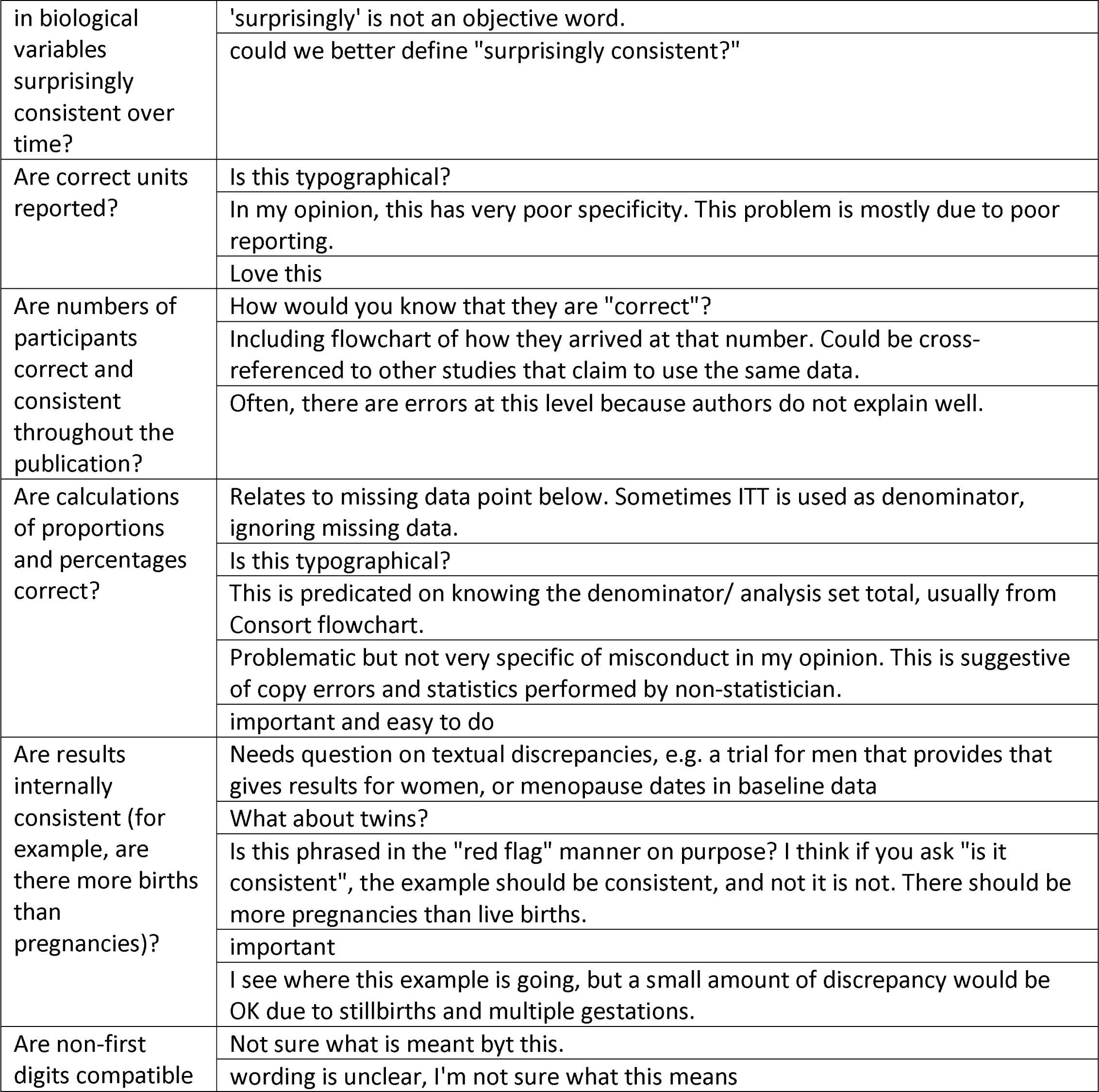
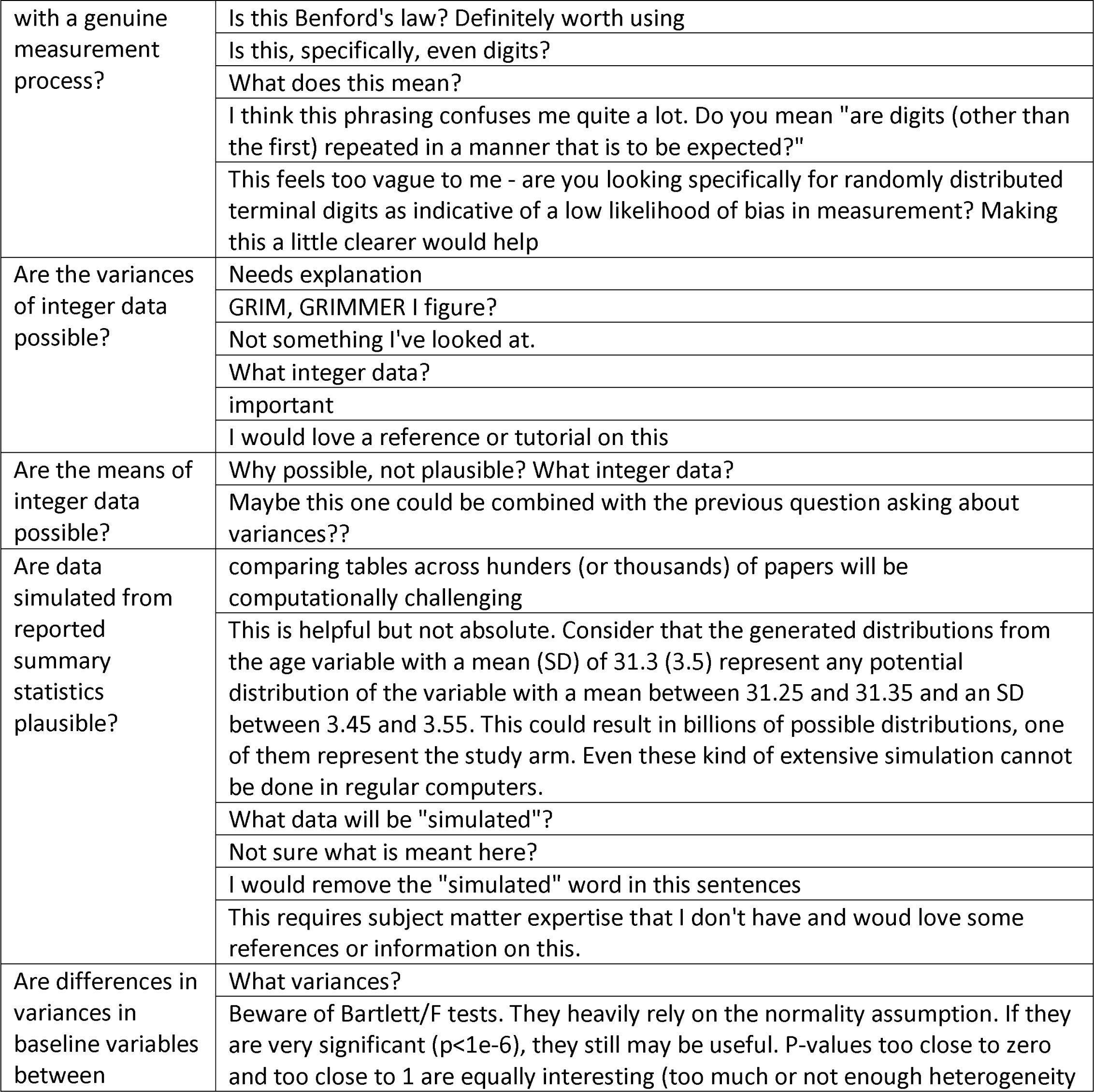
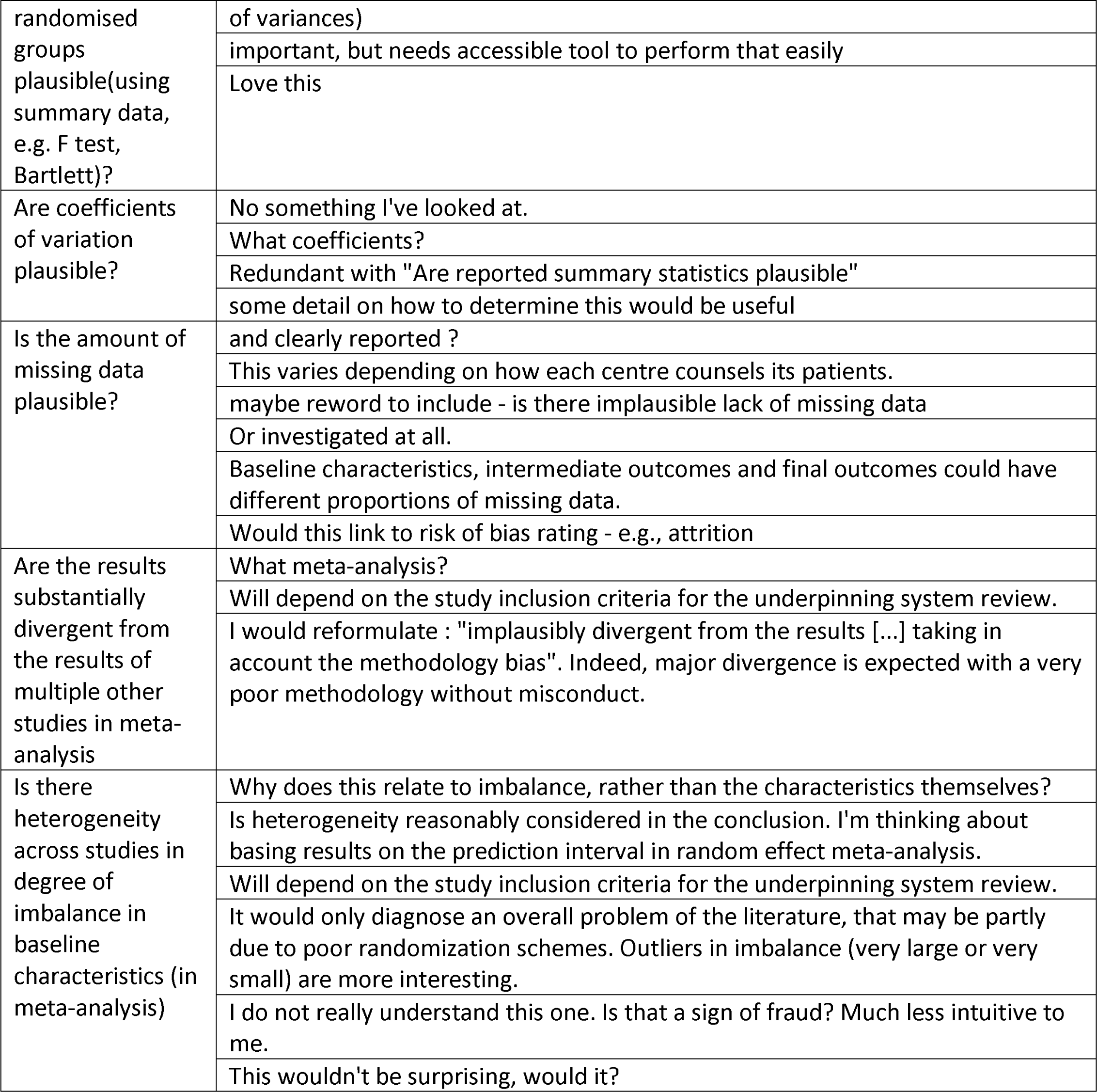

#### Inspecting the research team

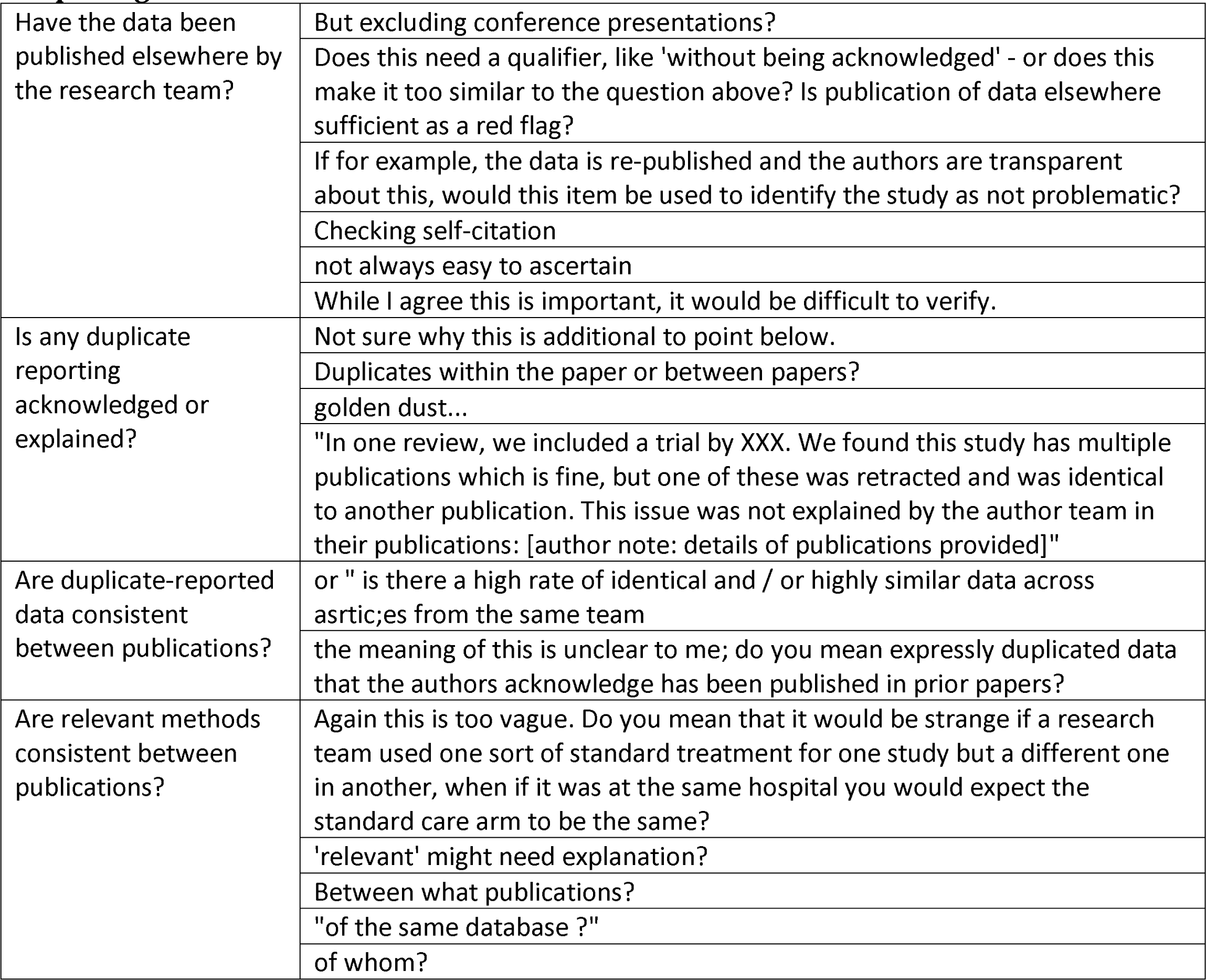
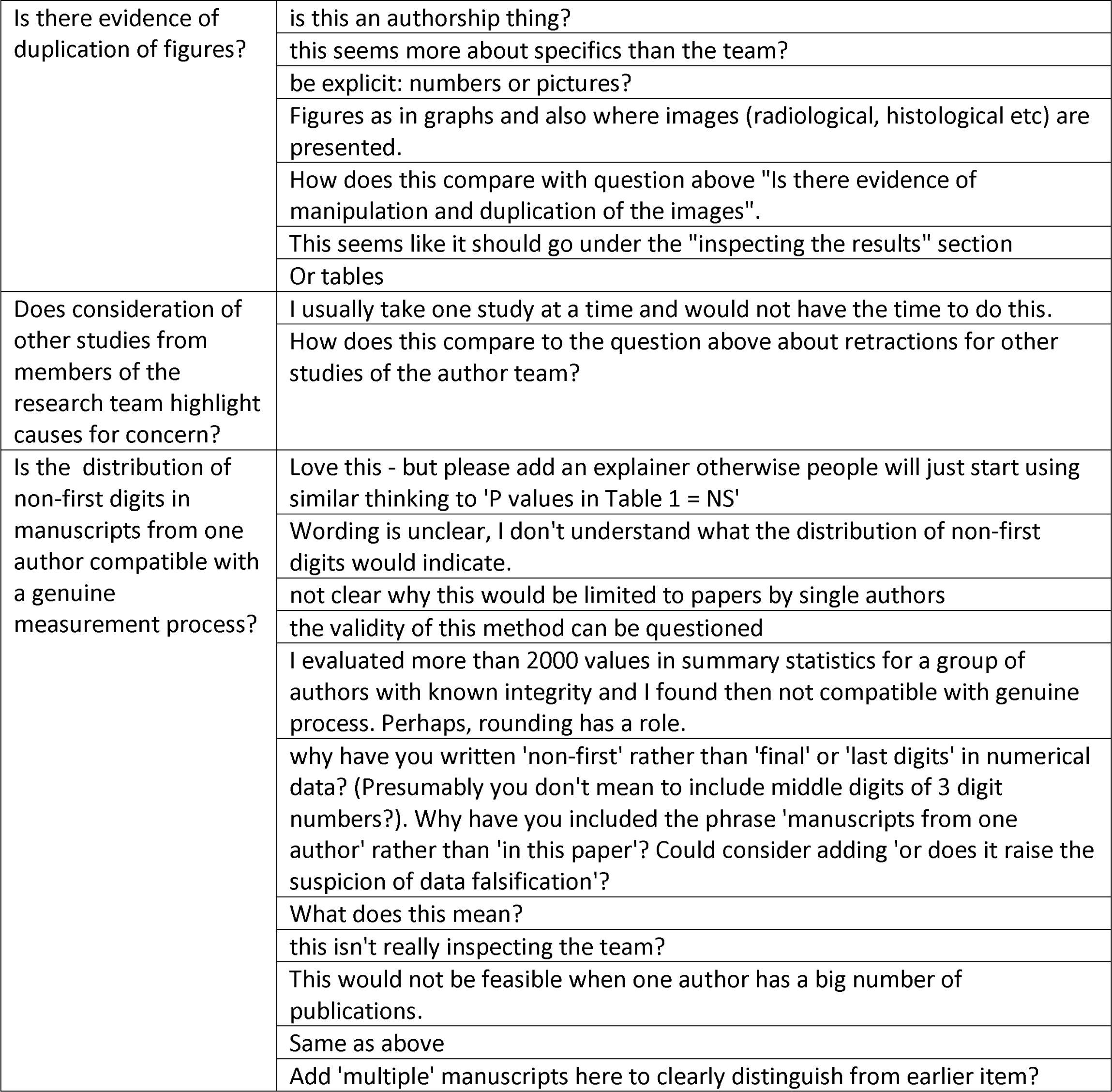
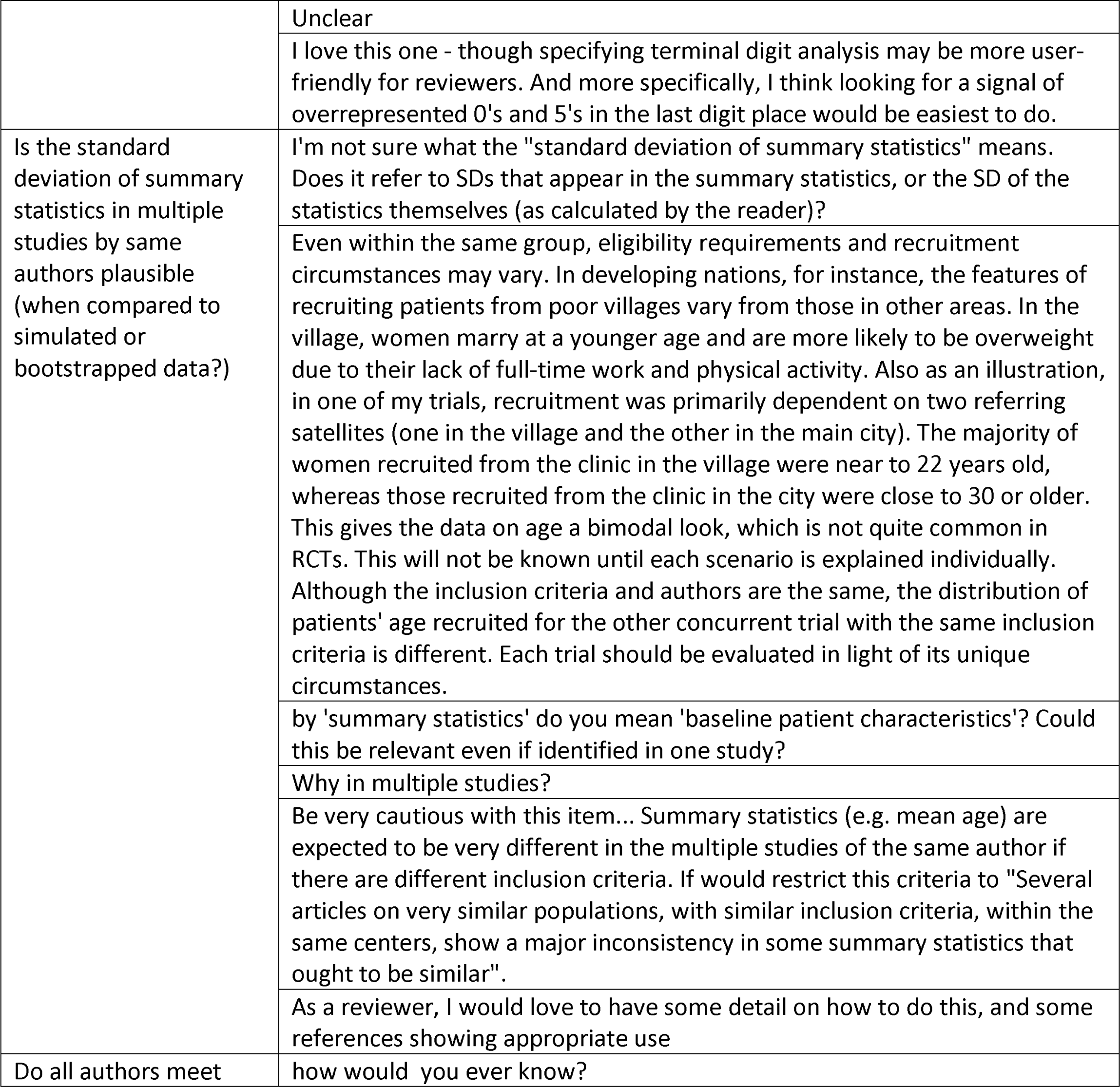
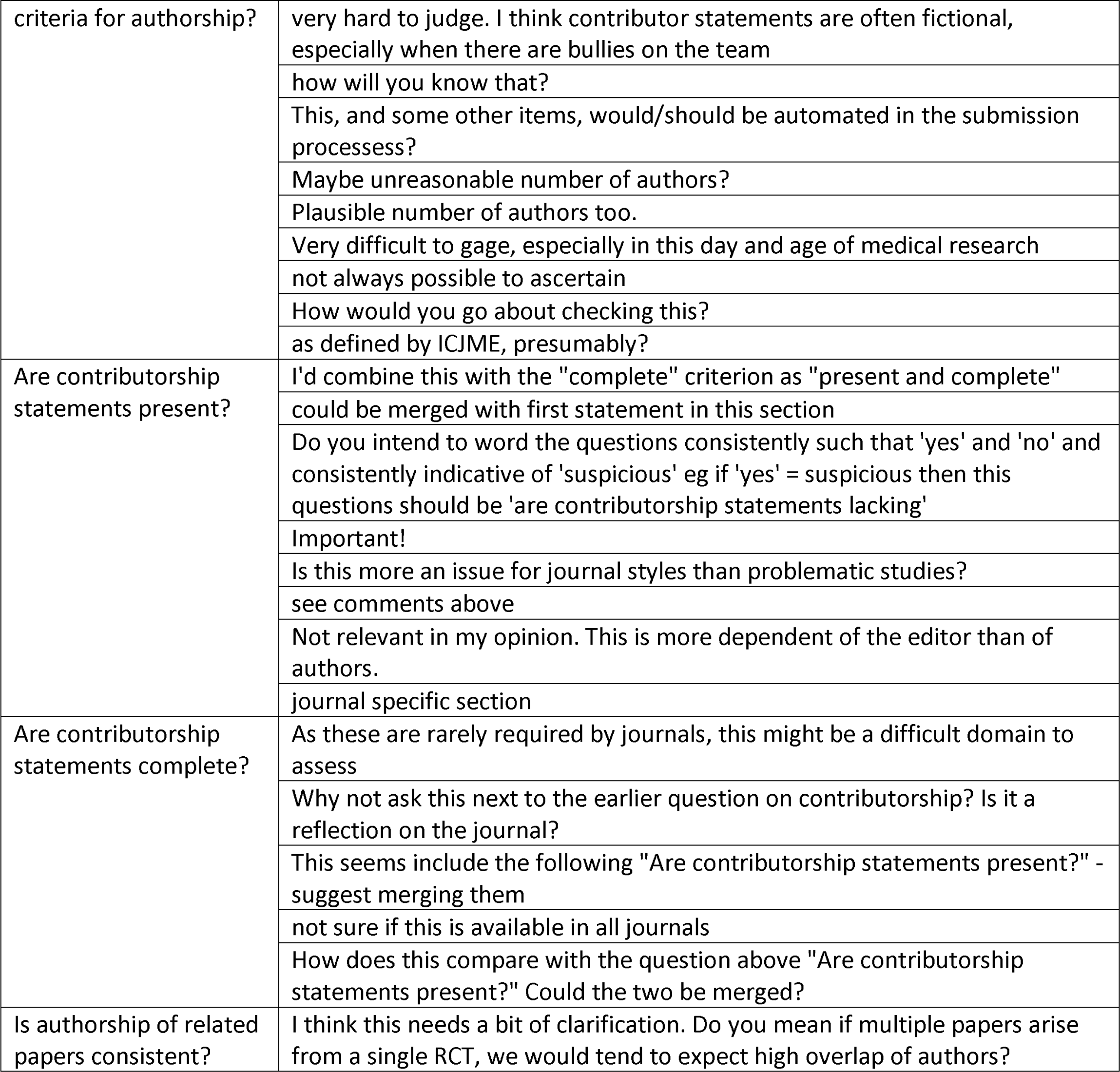
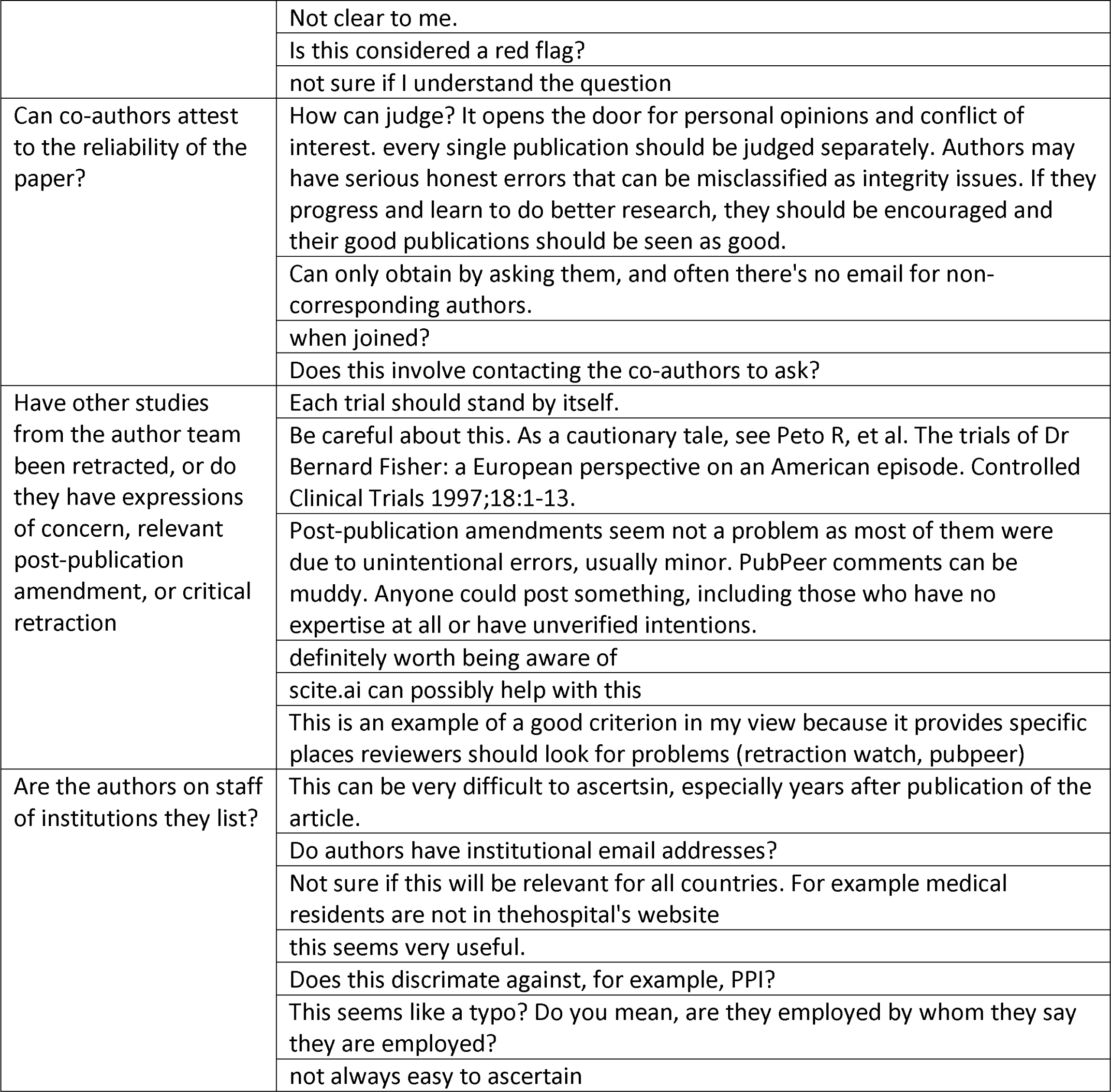
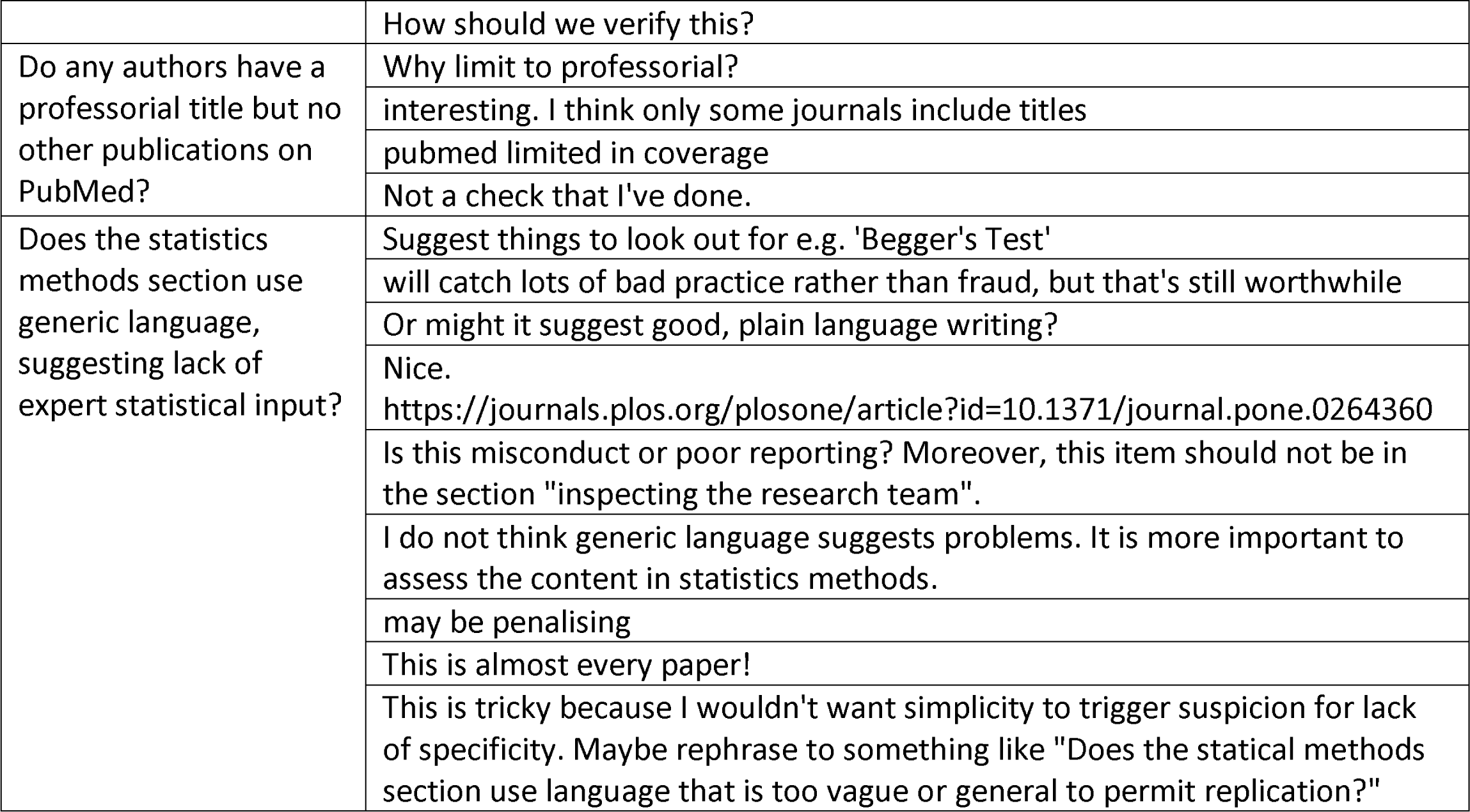

#### Inspecting conduct, governance and transparency

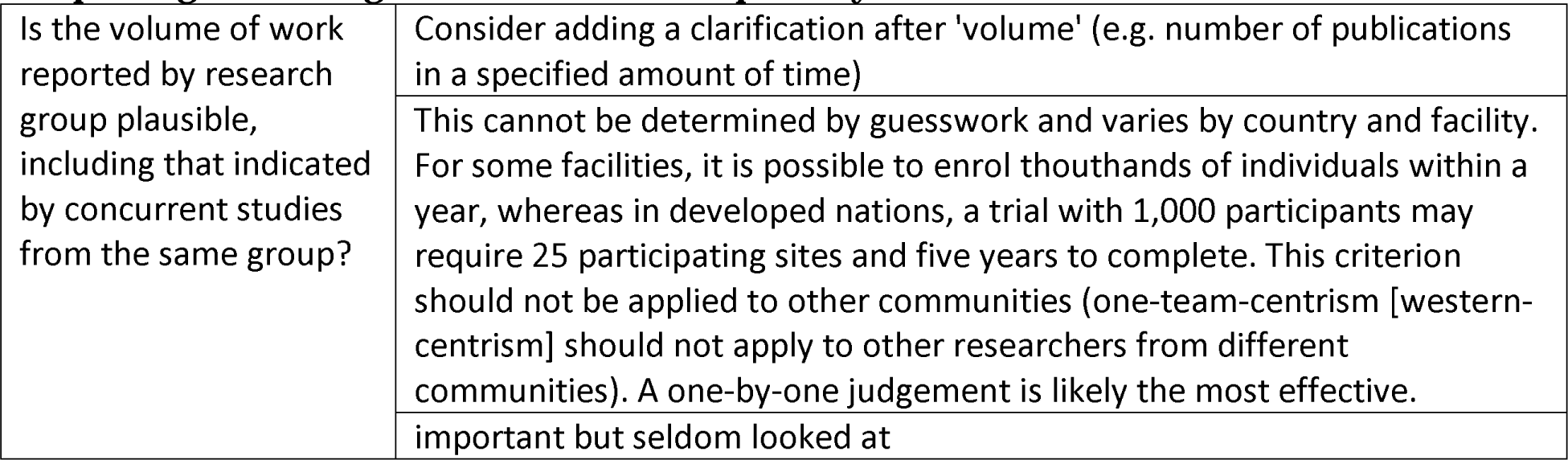
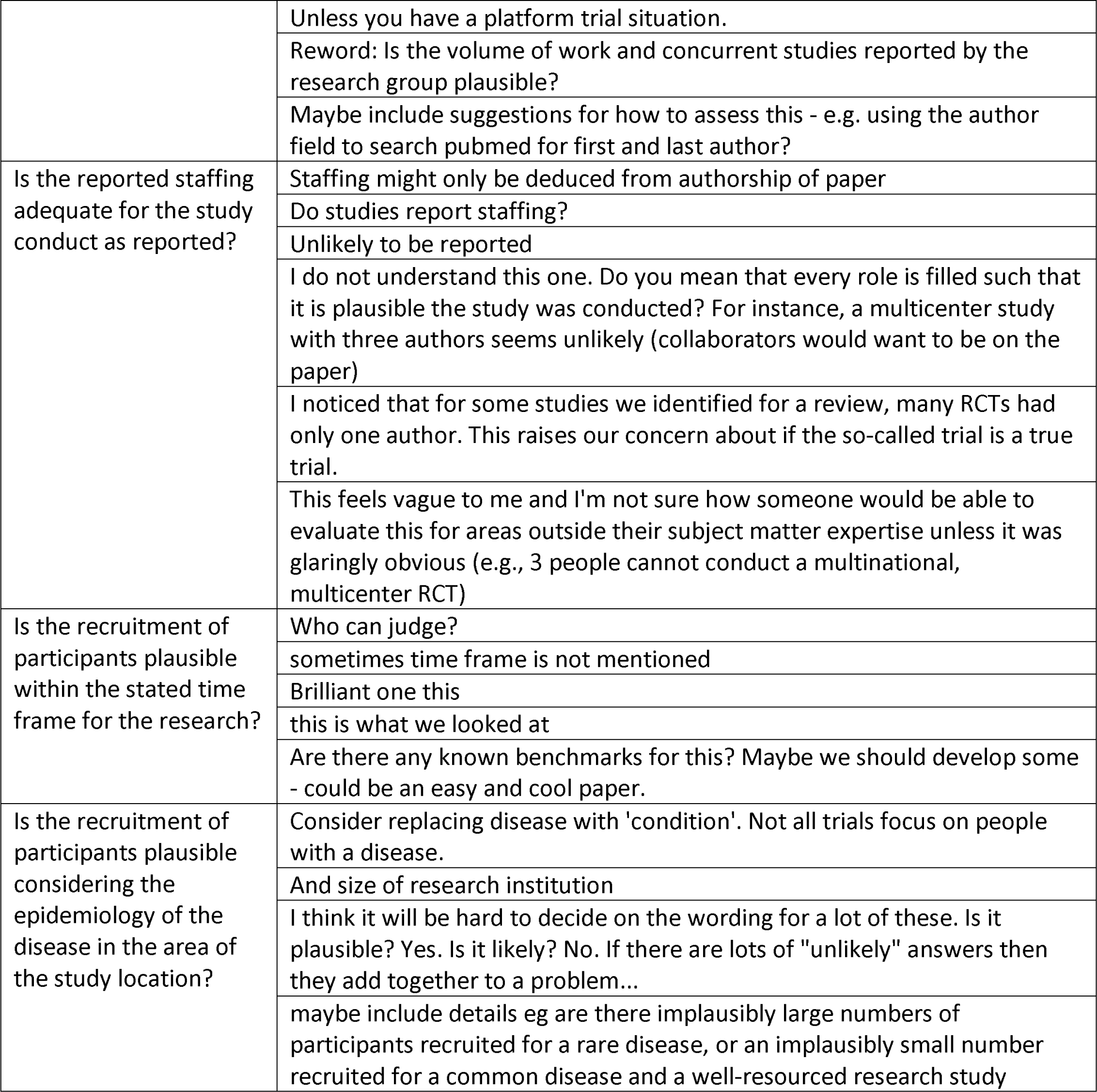
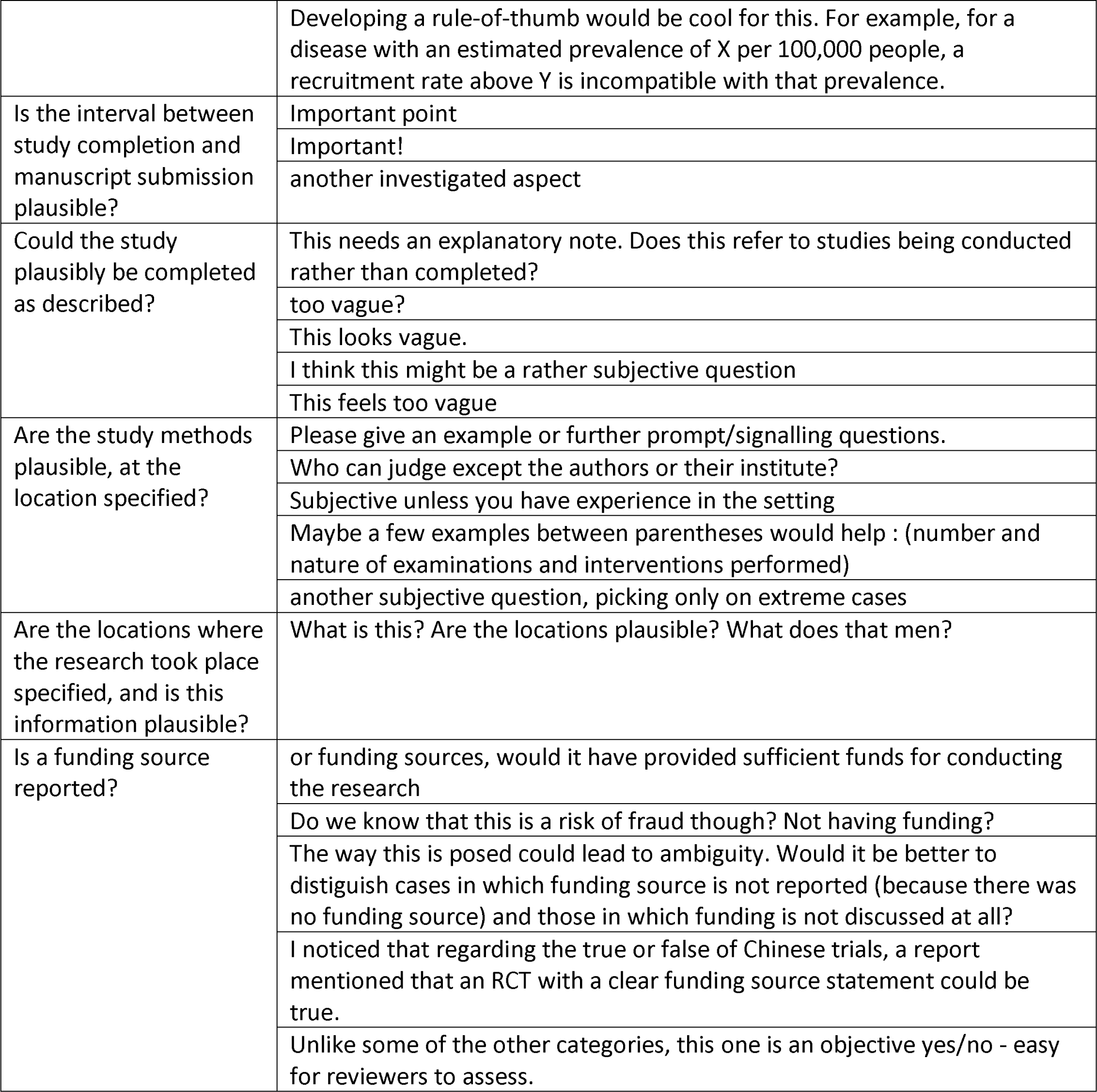
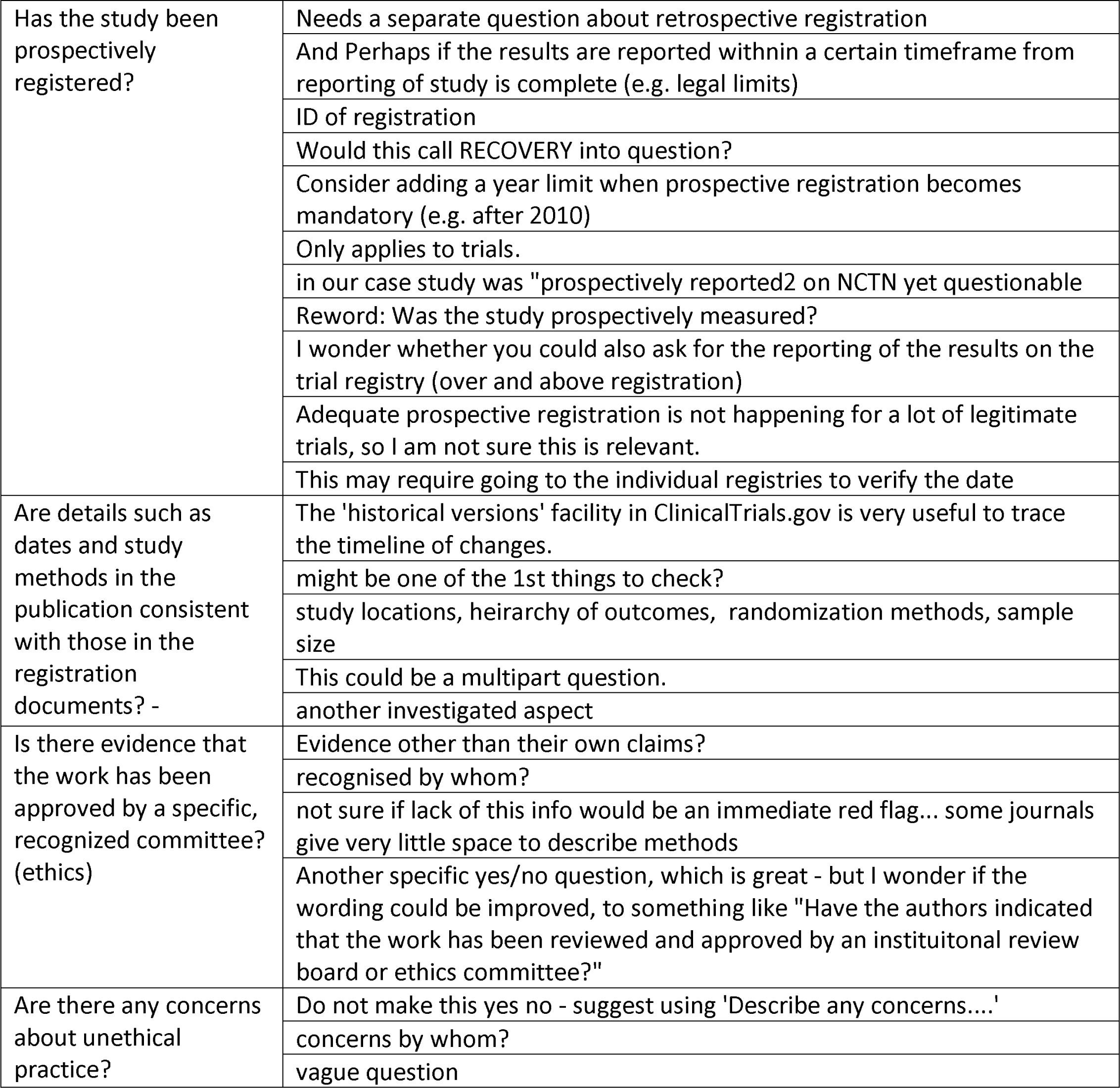
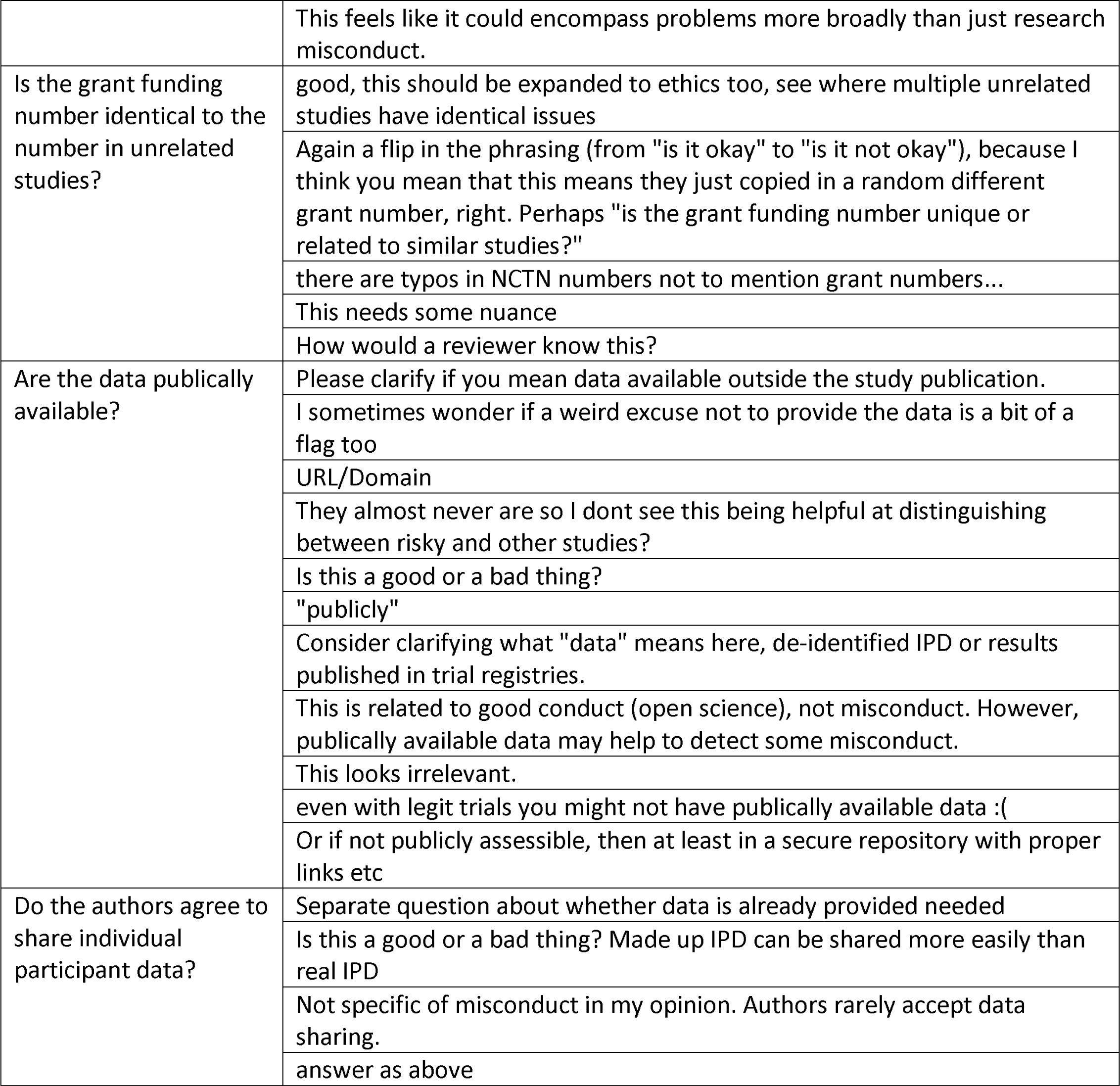
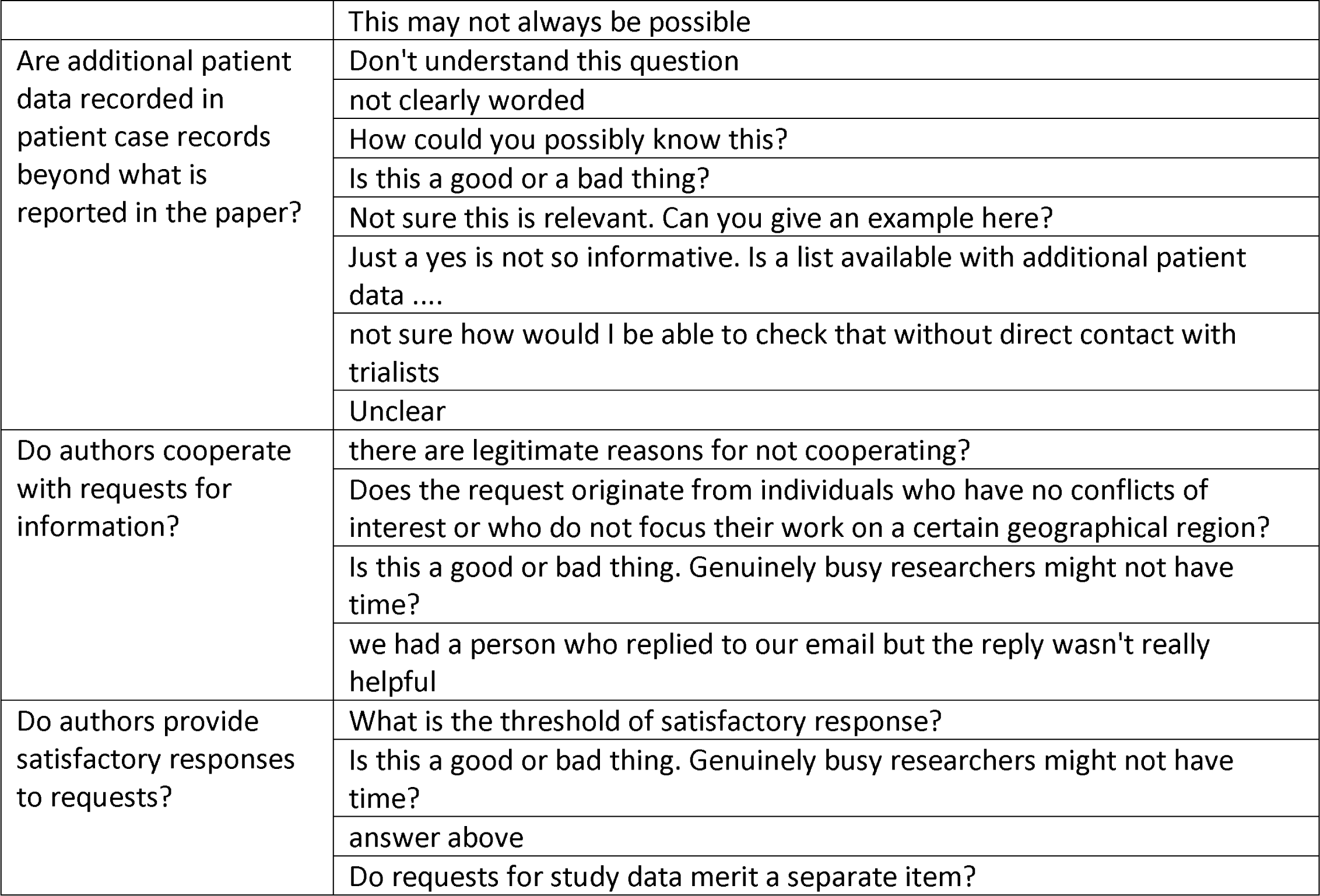

#### Inspecting text and publication details

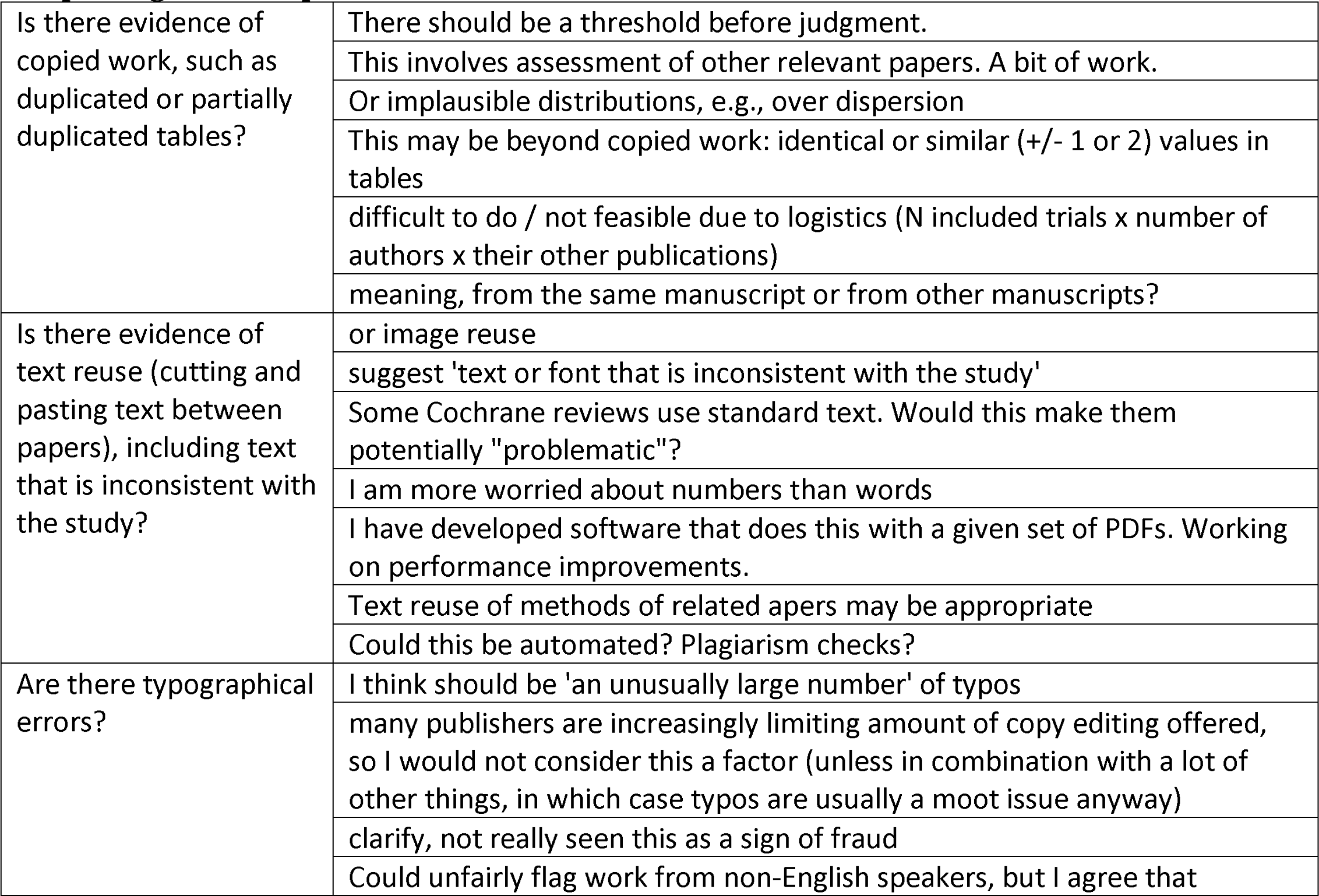
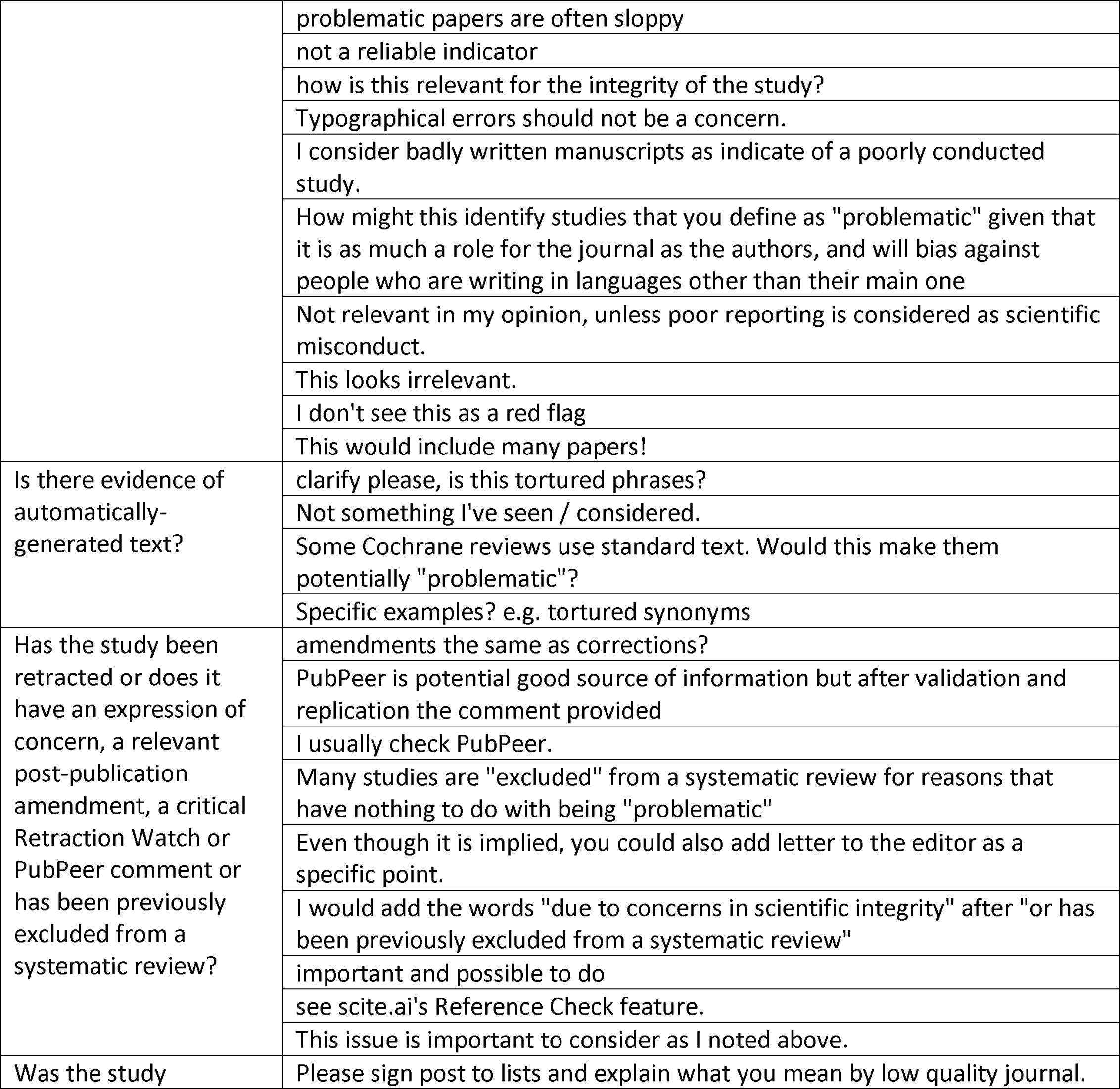
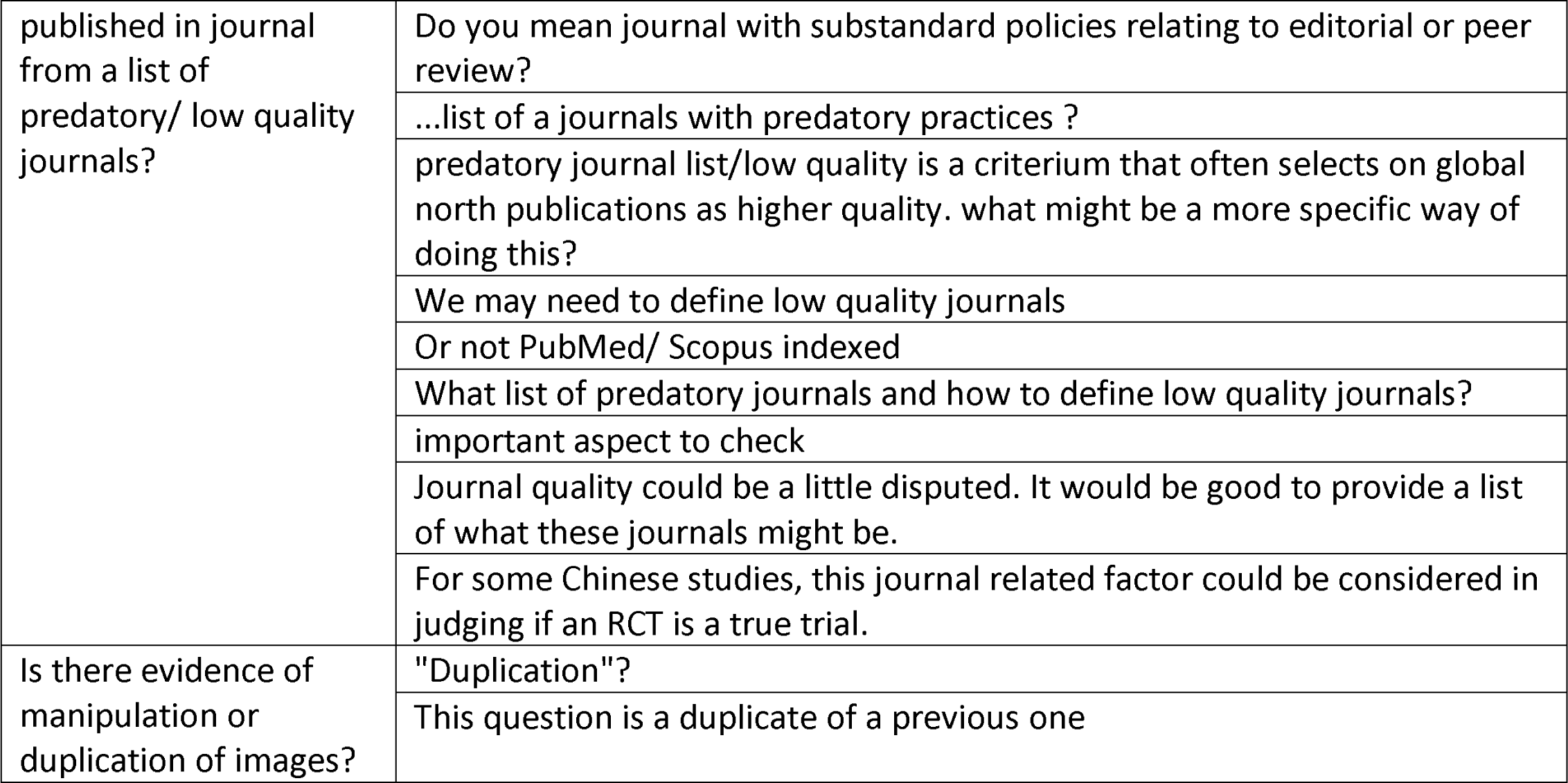

#### Inspecting individual participant data

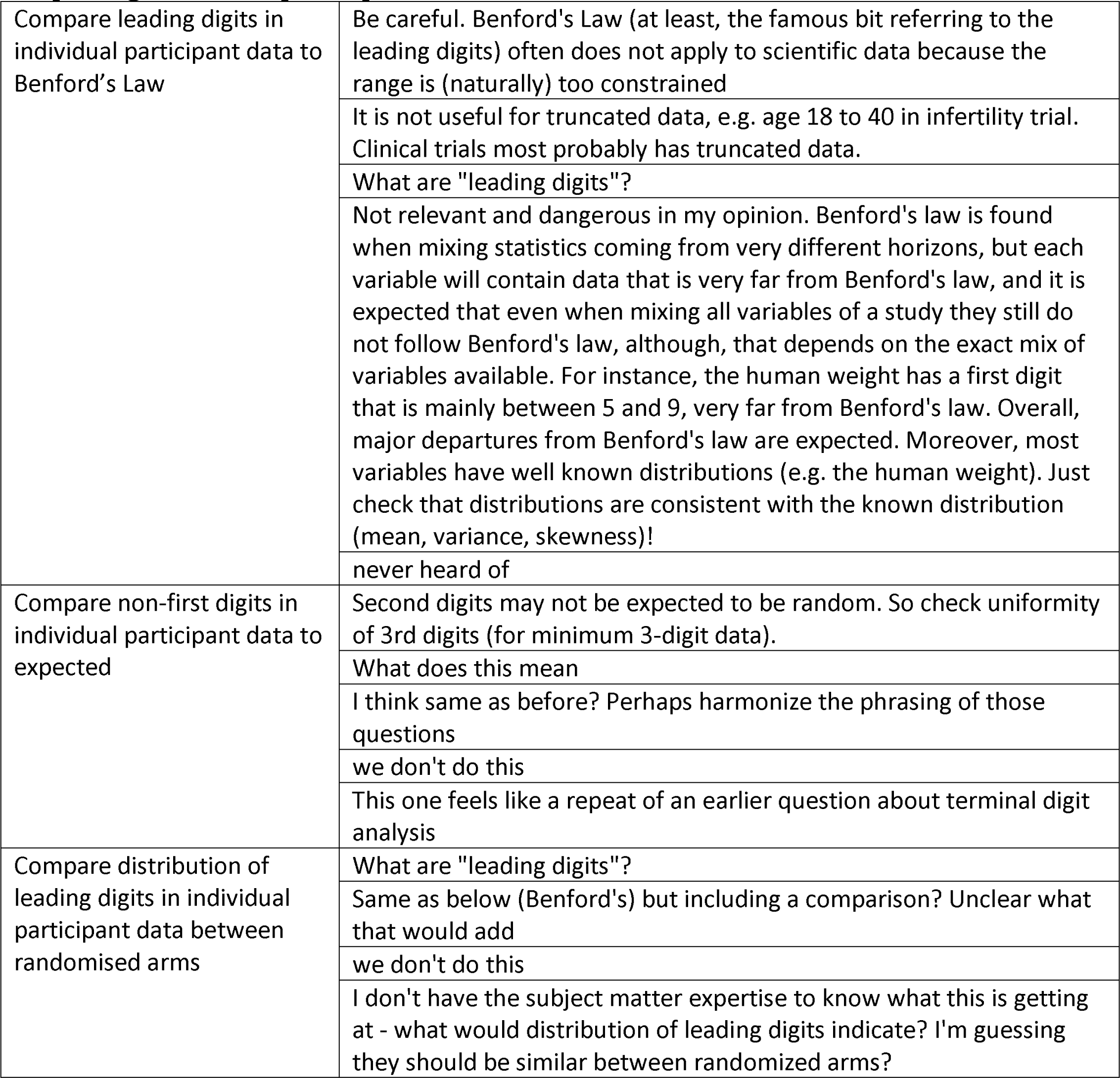
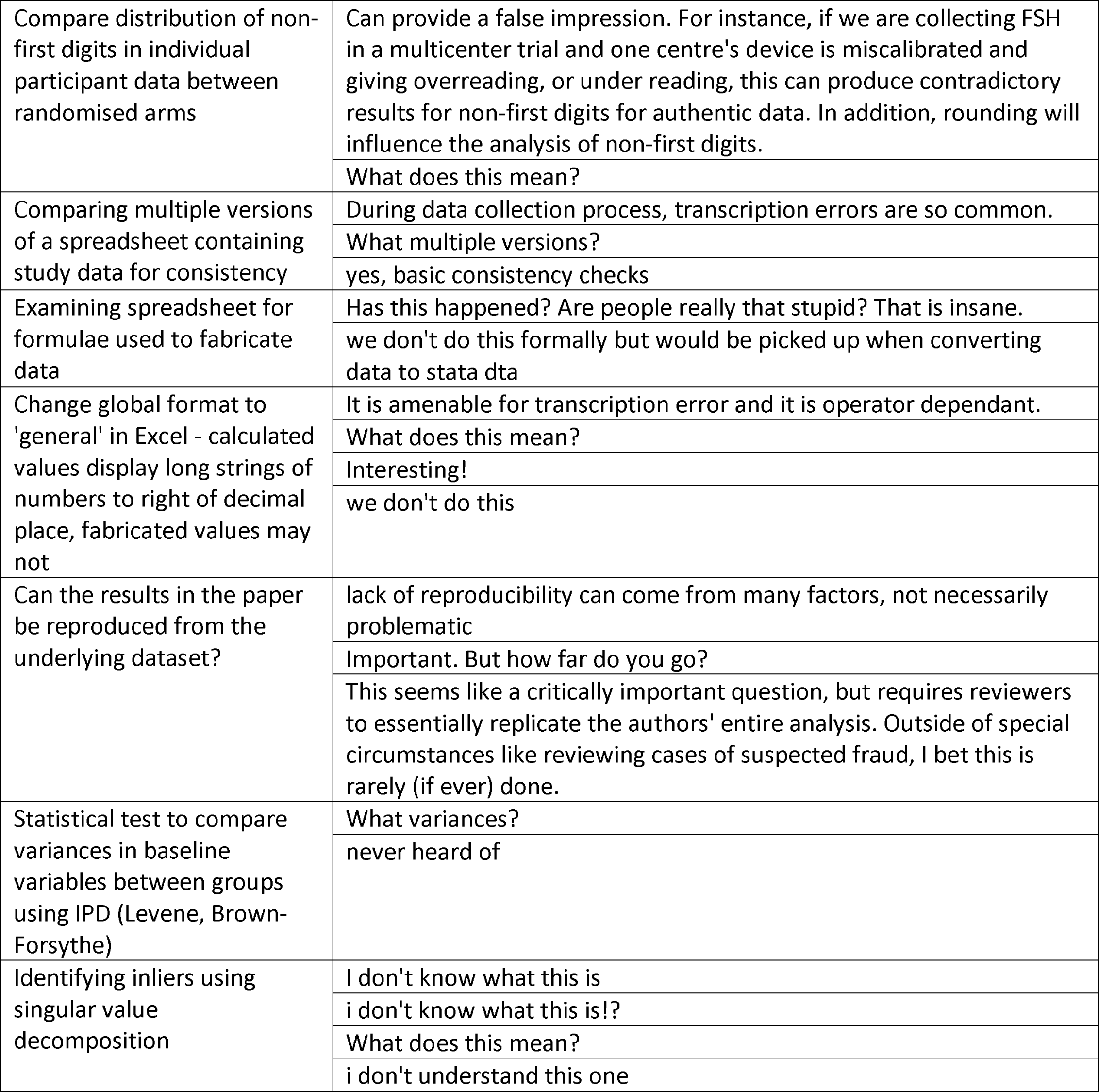
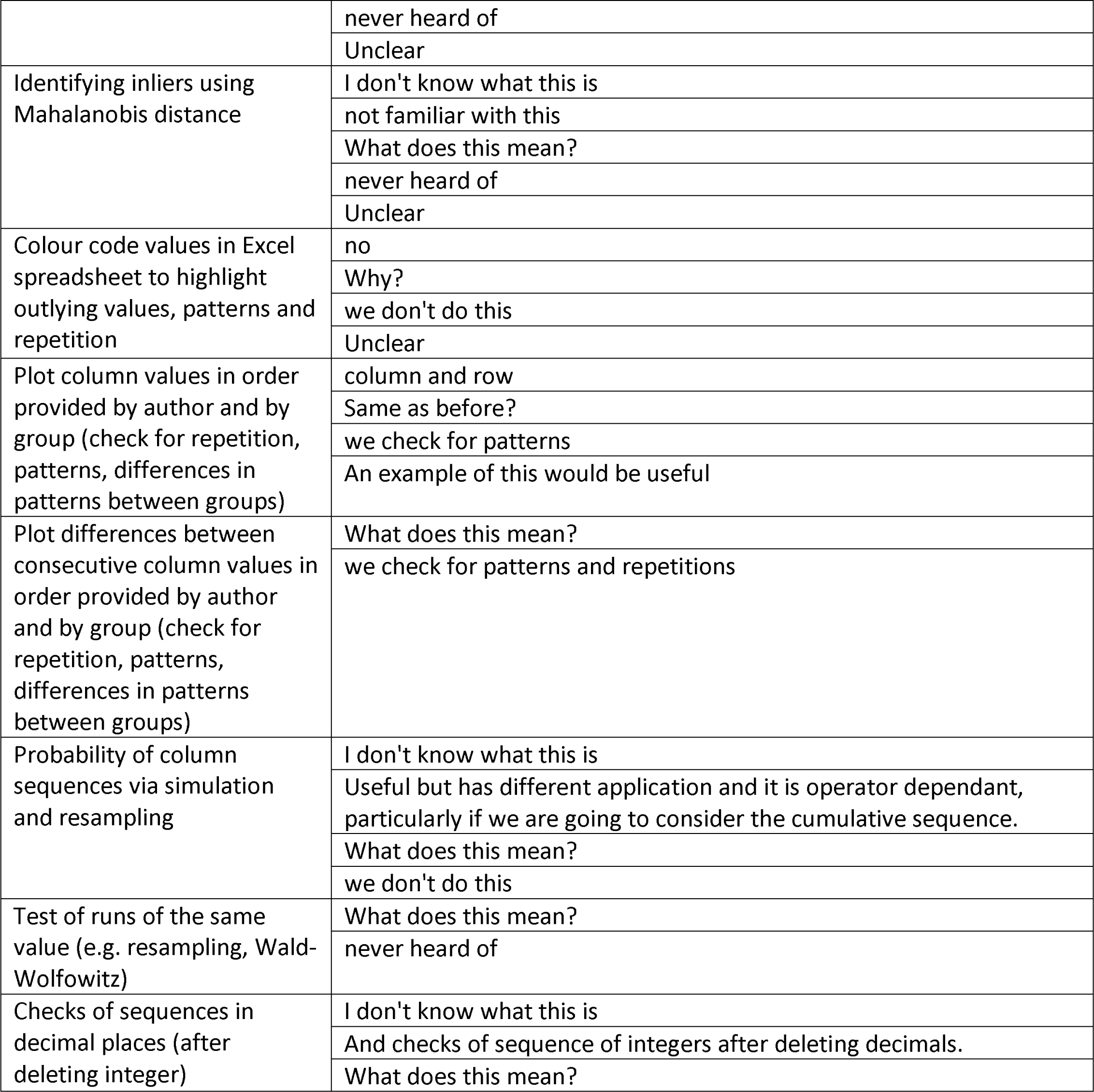
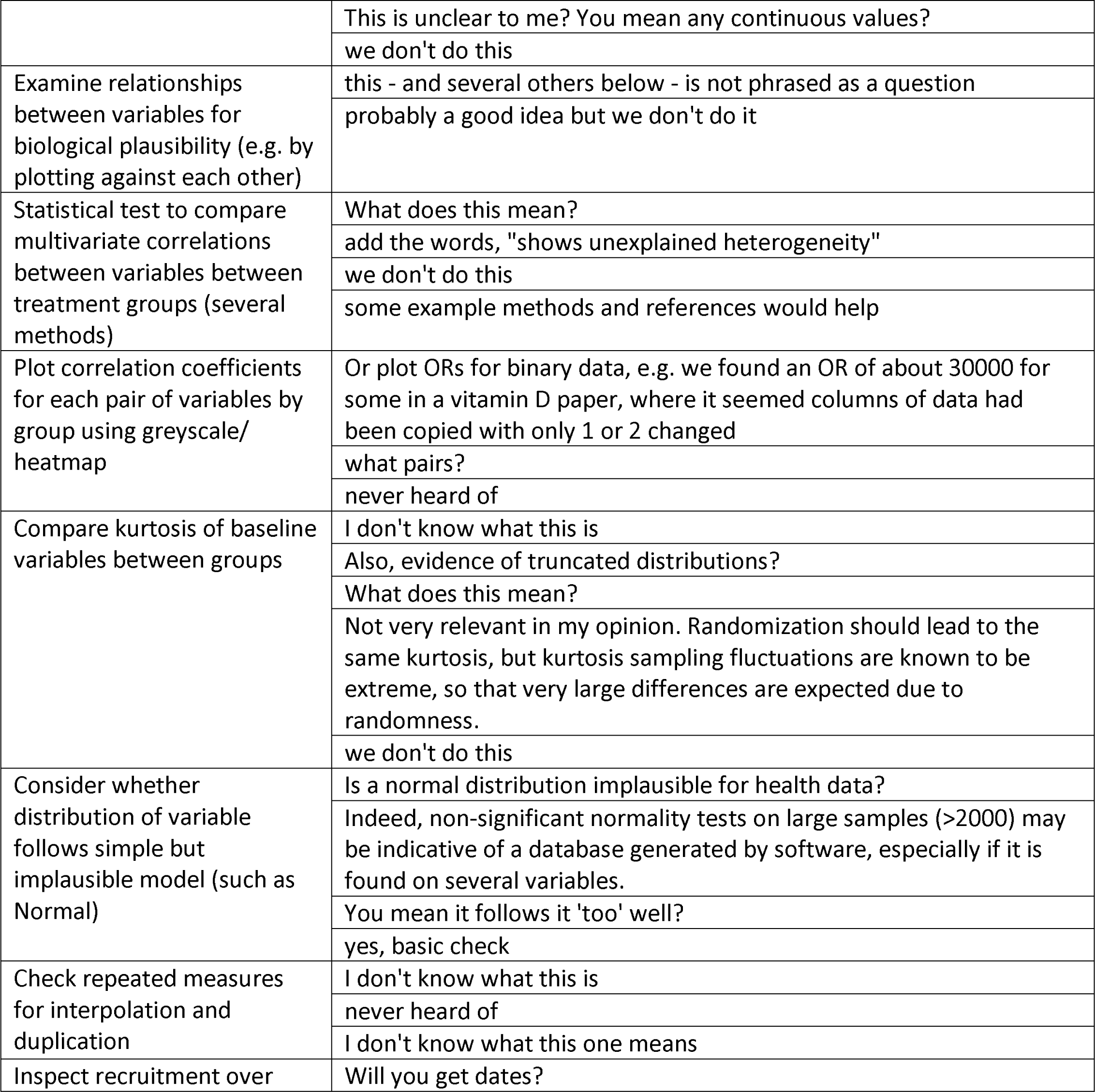
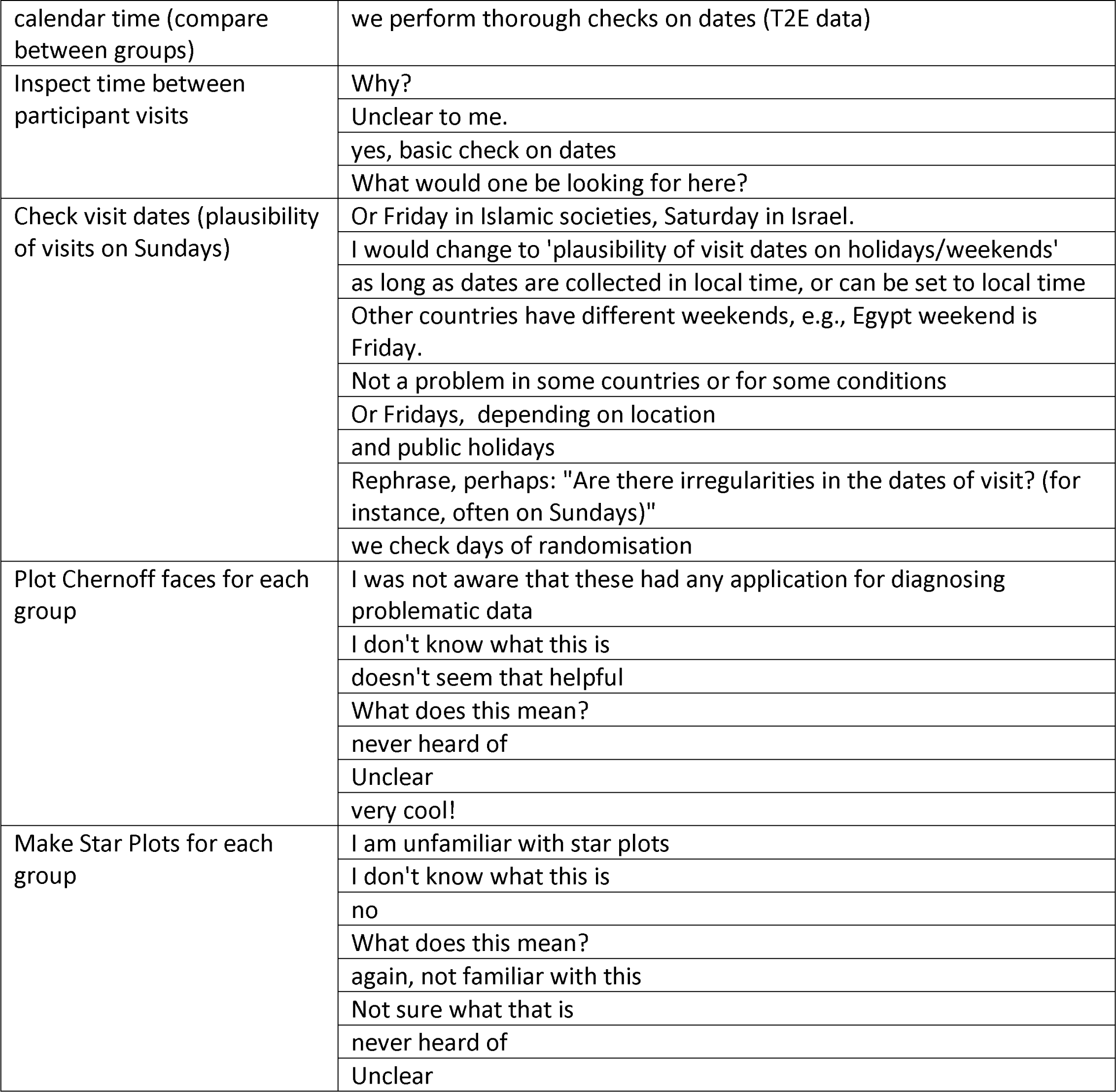
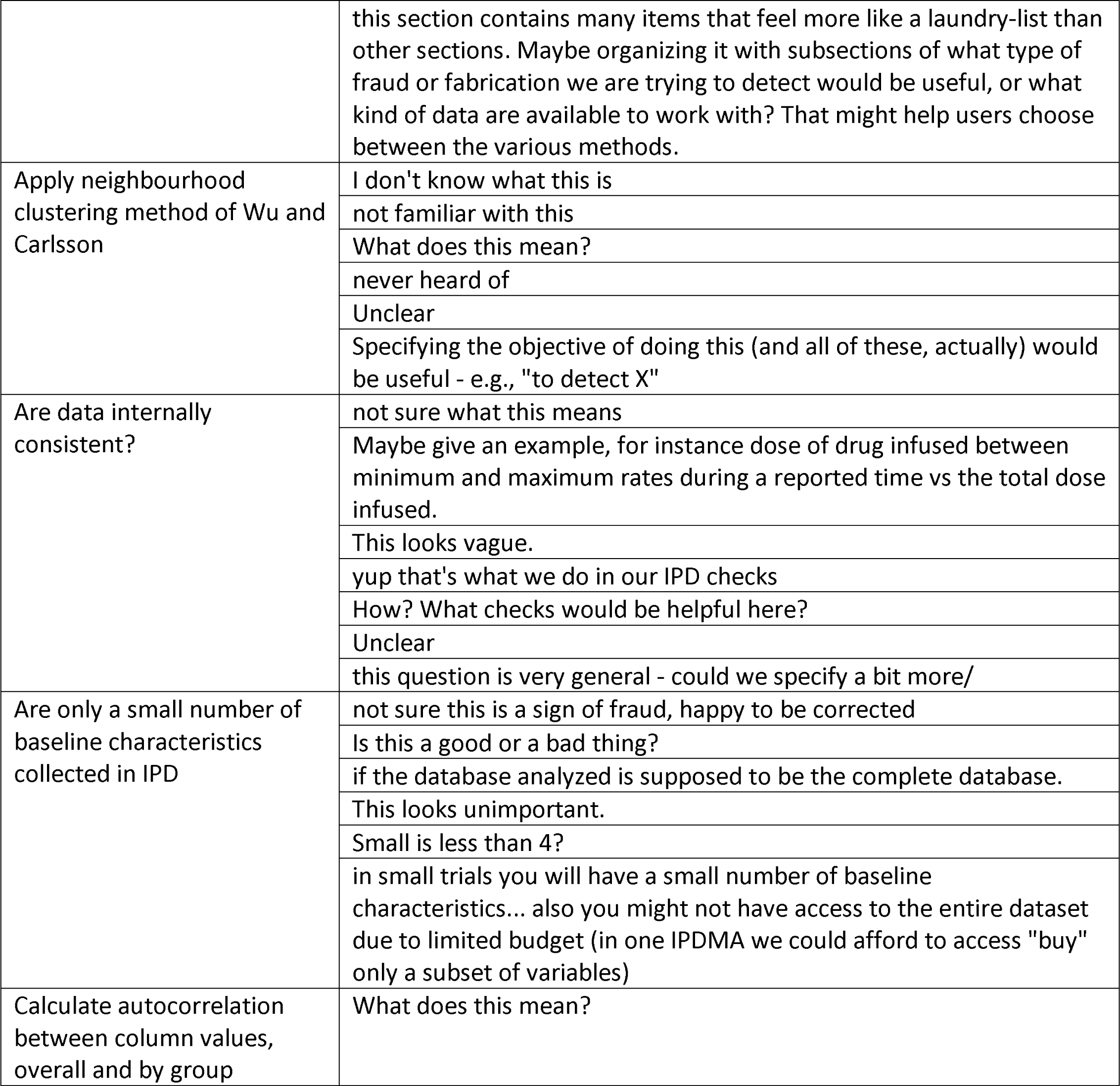
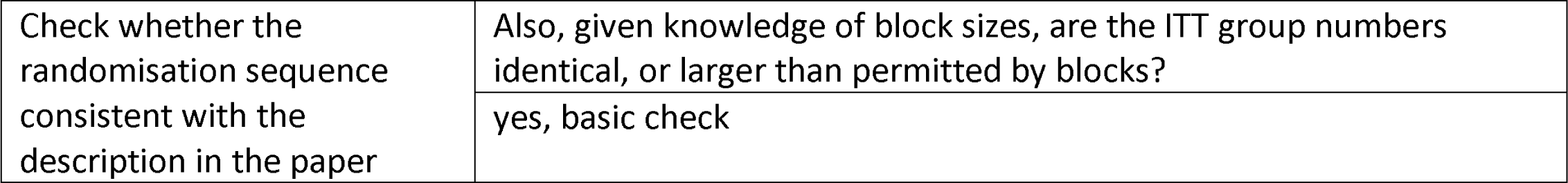

#### Do you know of any methods not listed here, or do you have any to suggest? Please describe them here if so

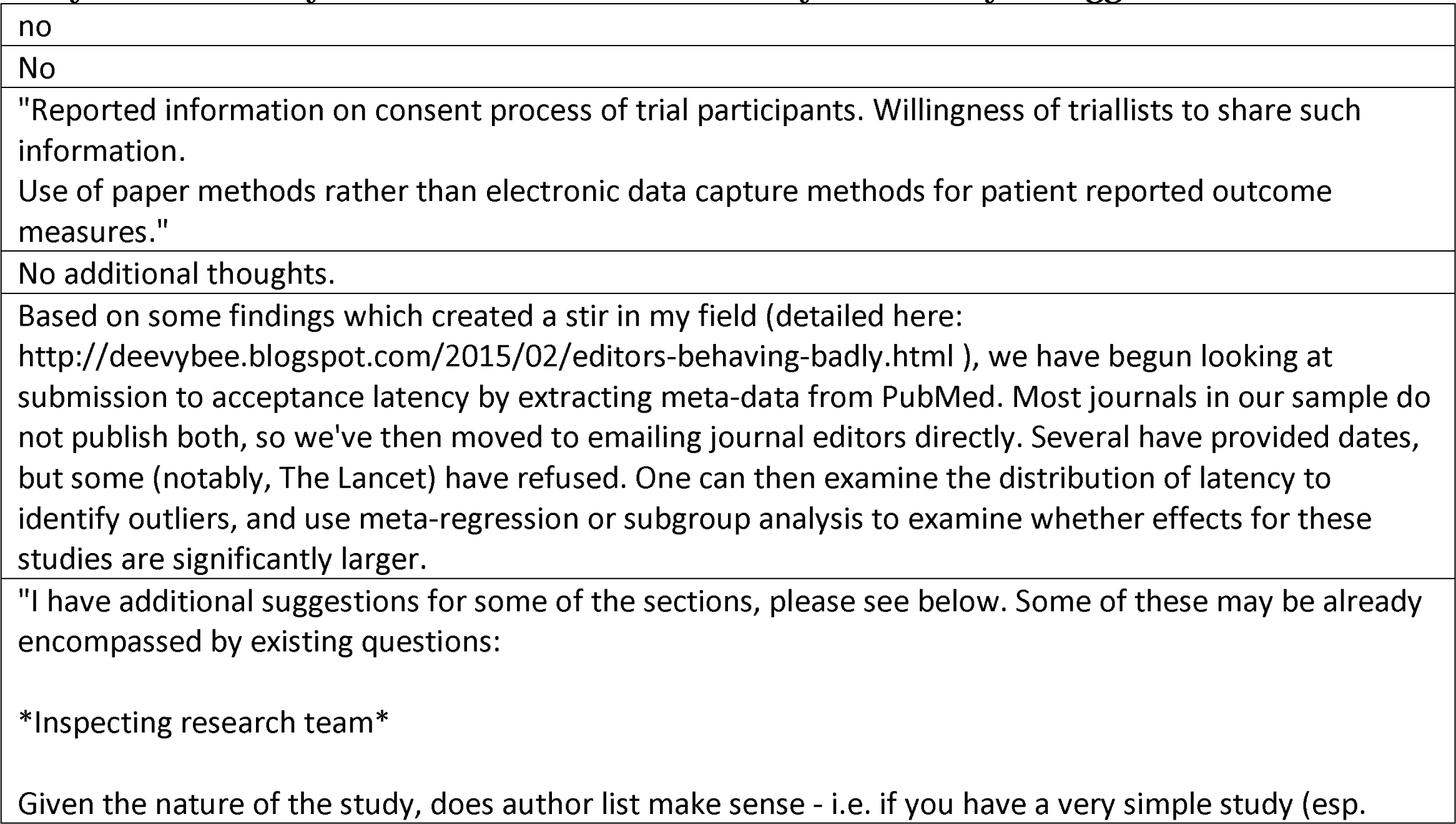
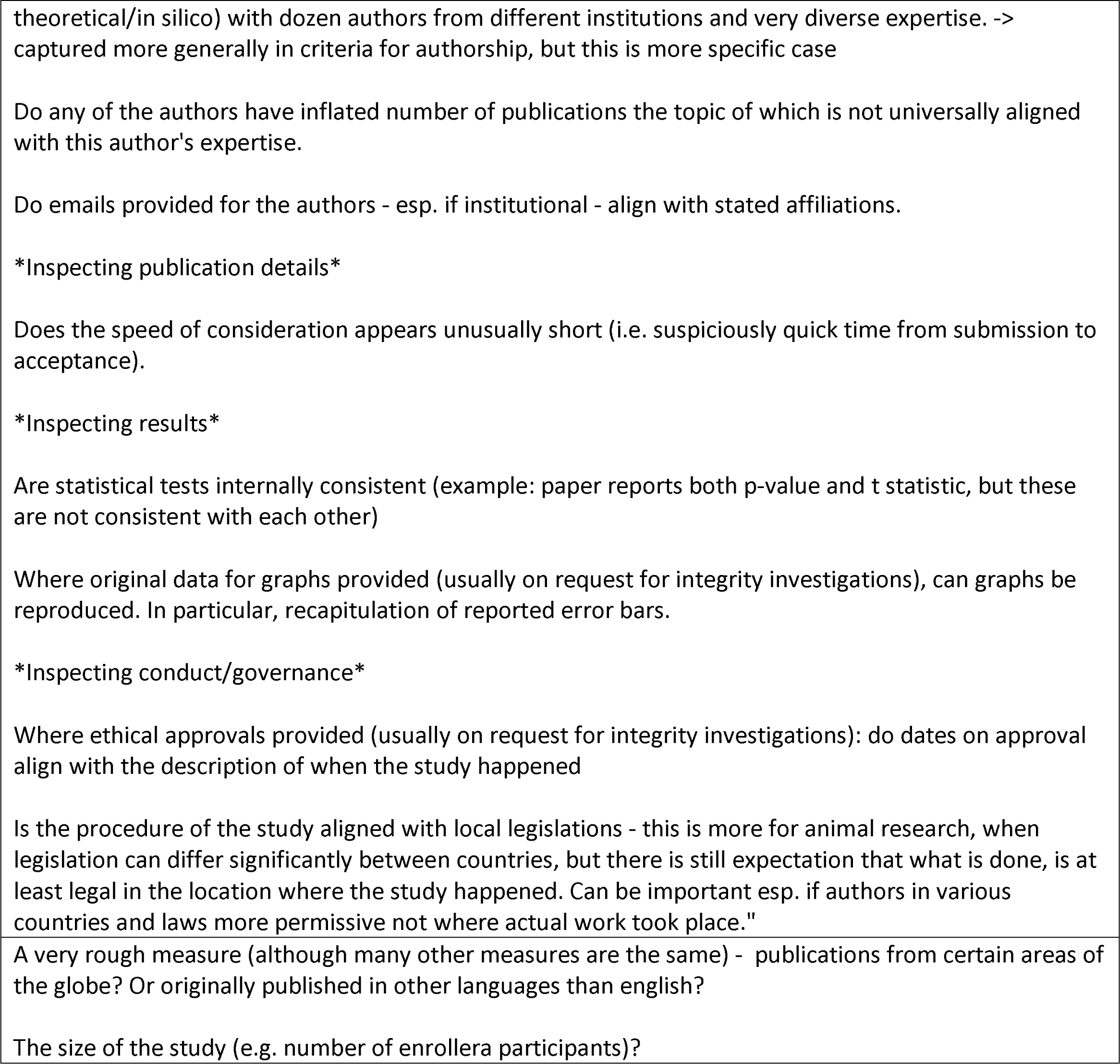
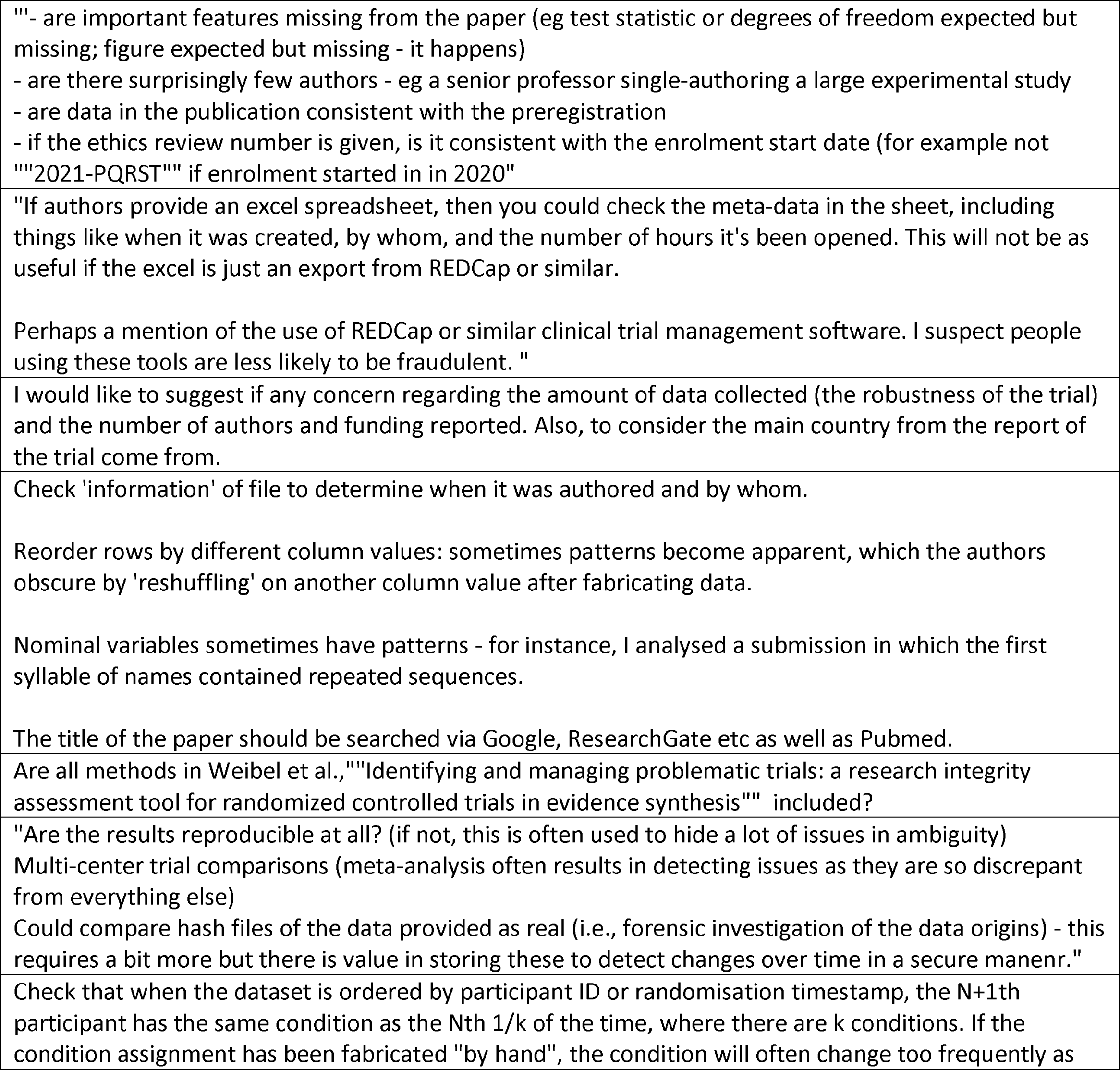
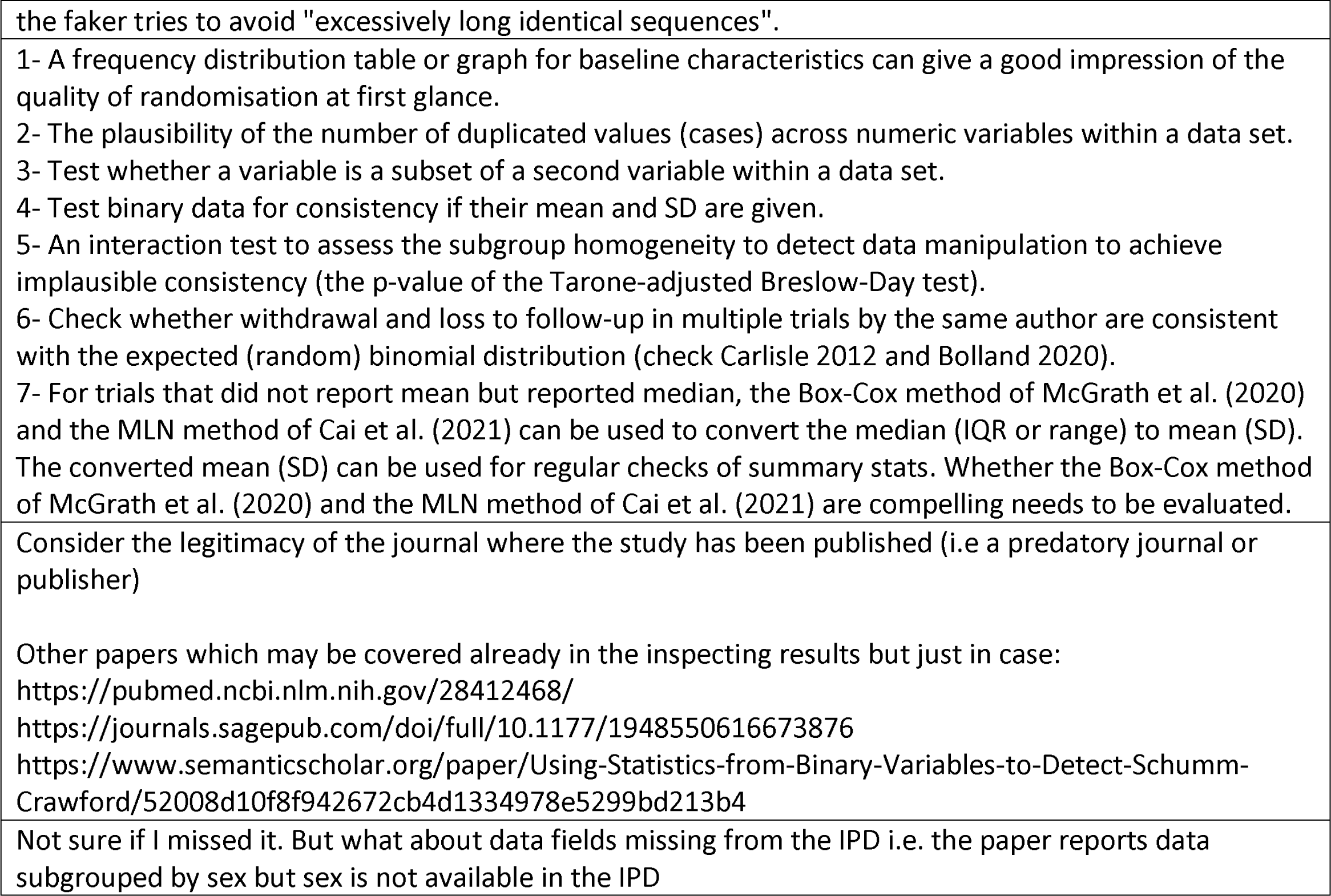

#### If you have any comments about this survey, or about this topic more generally, please add them here

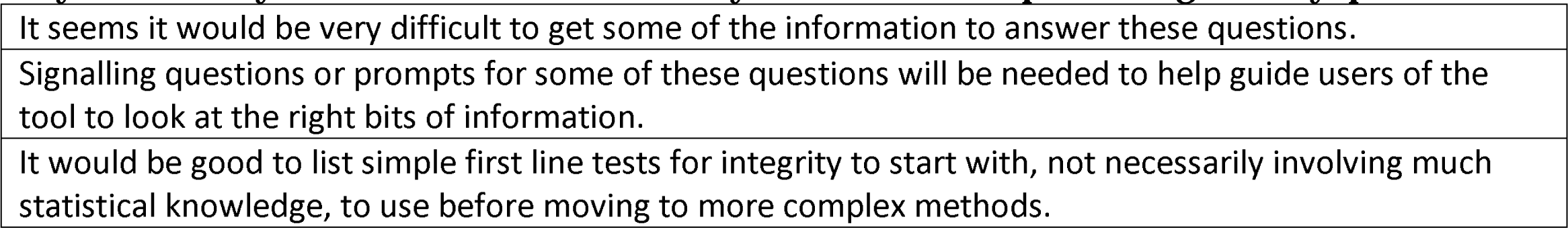
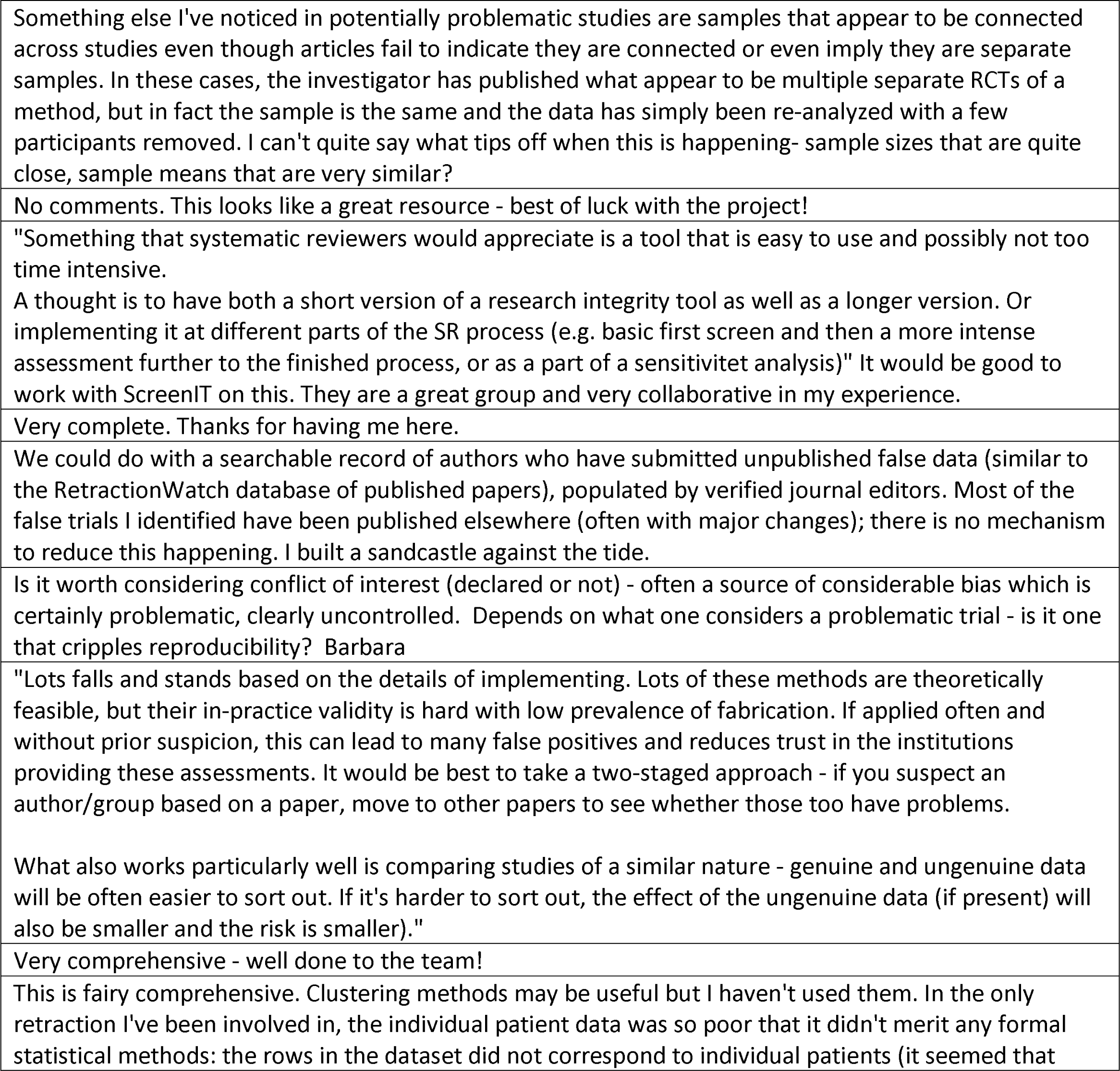
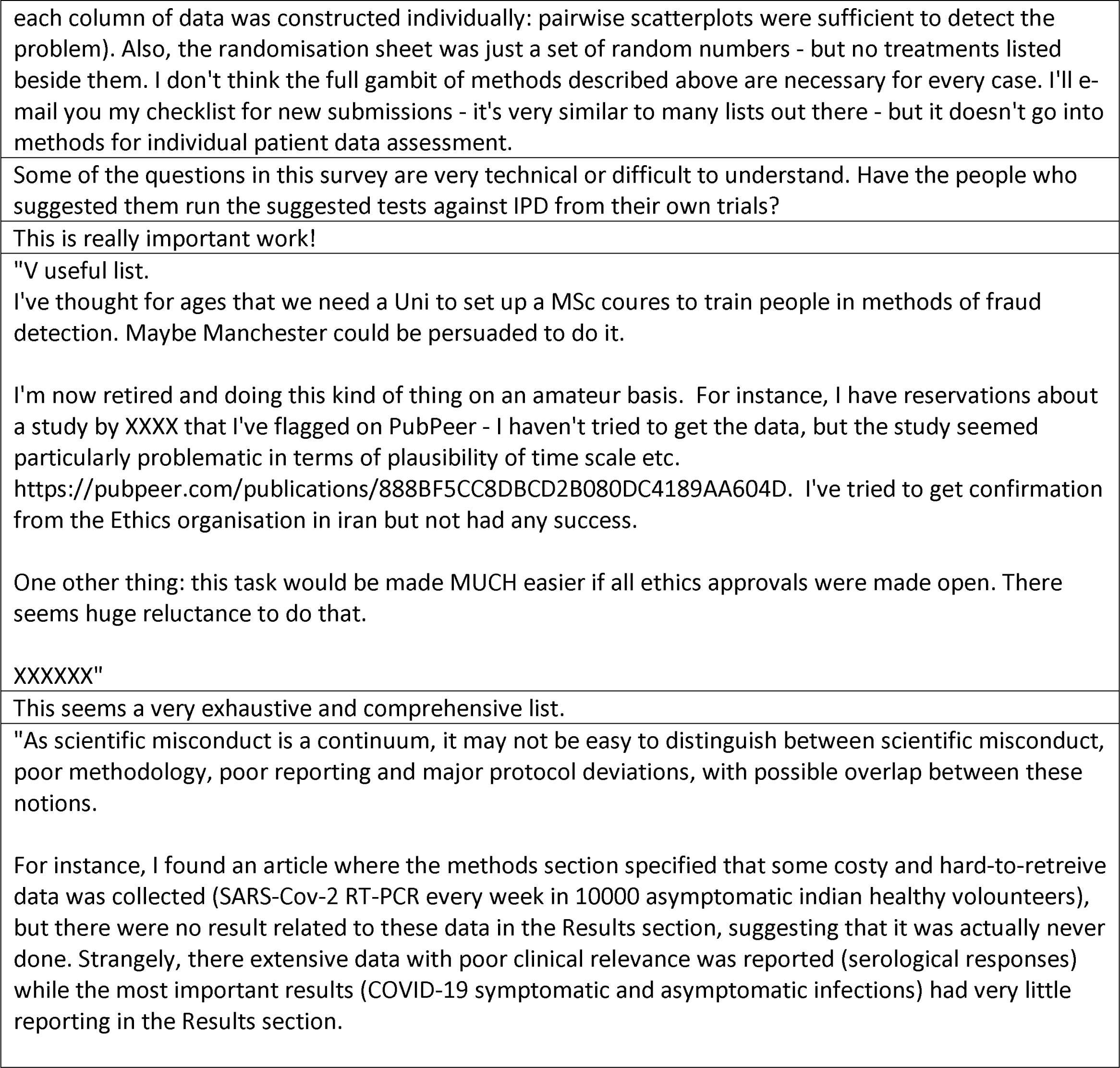
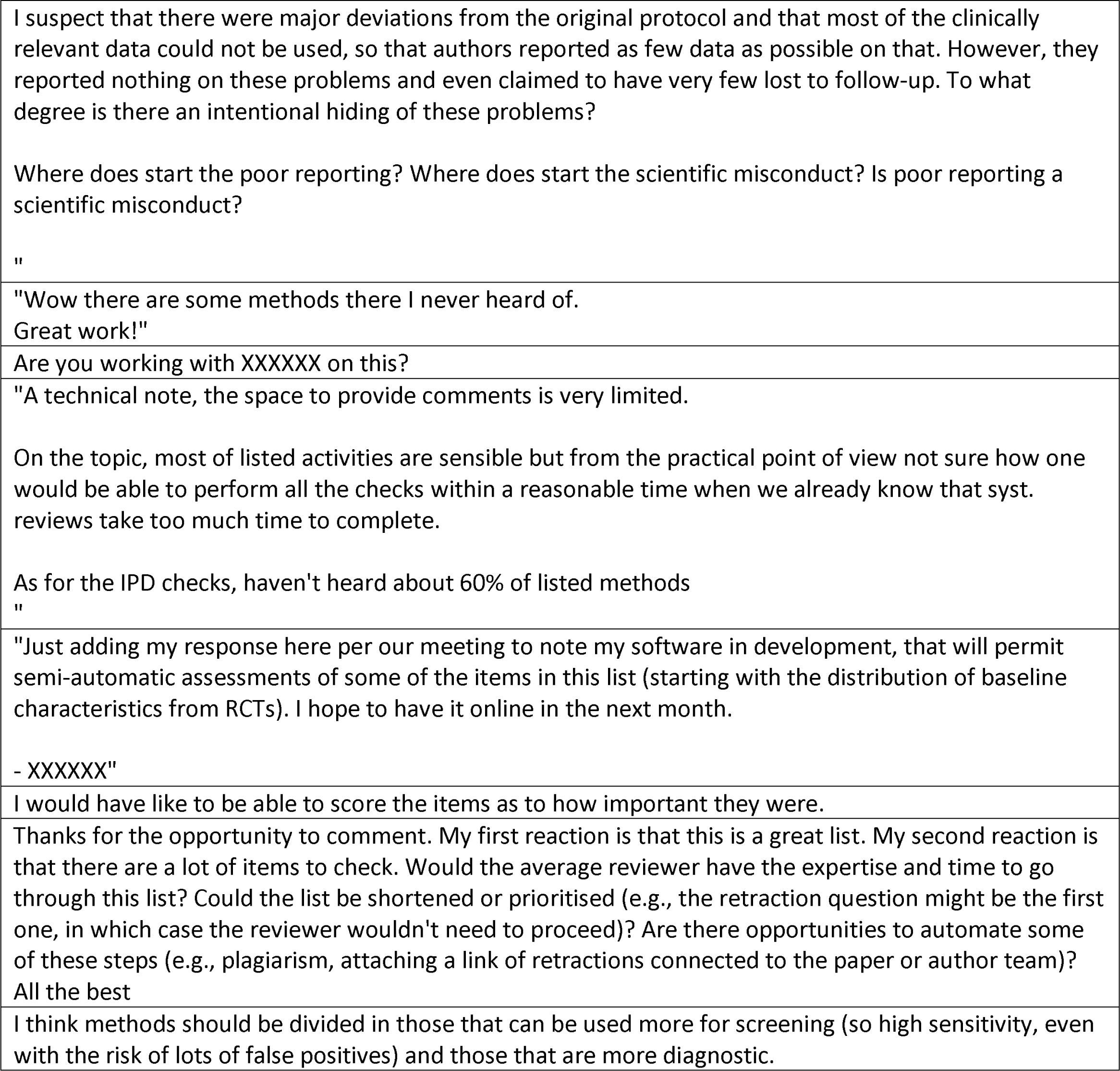
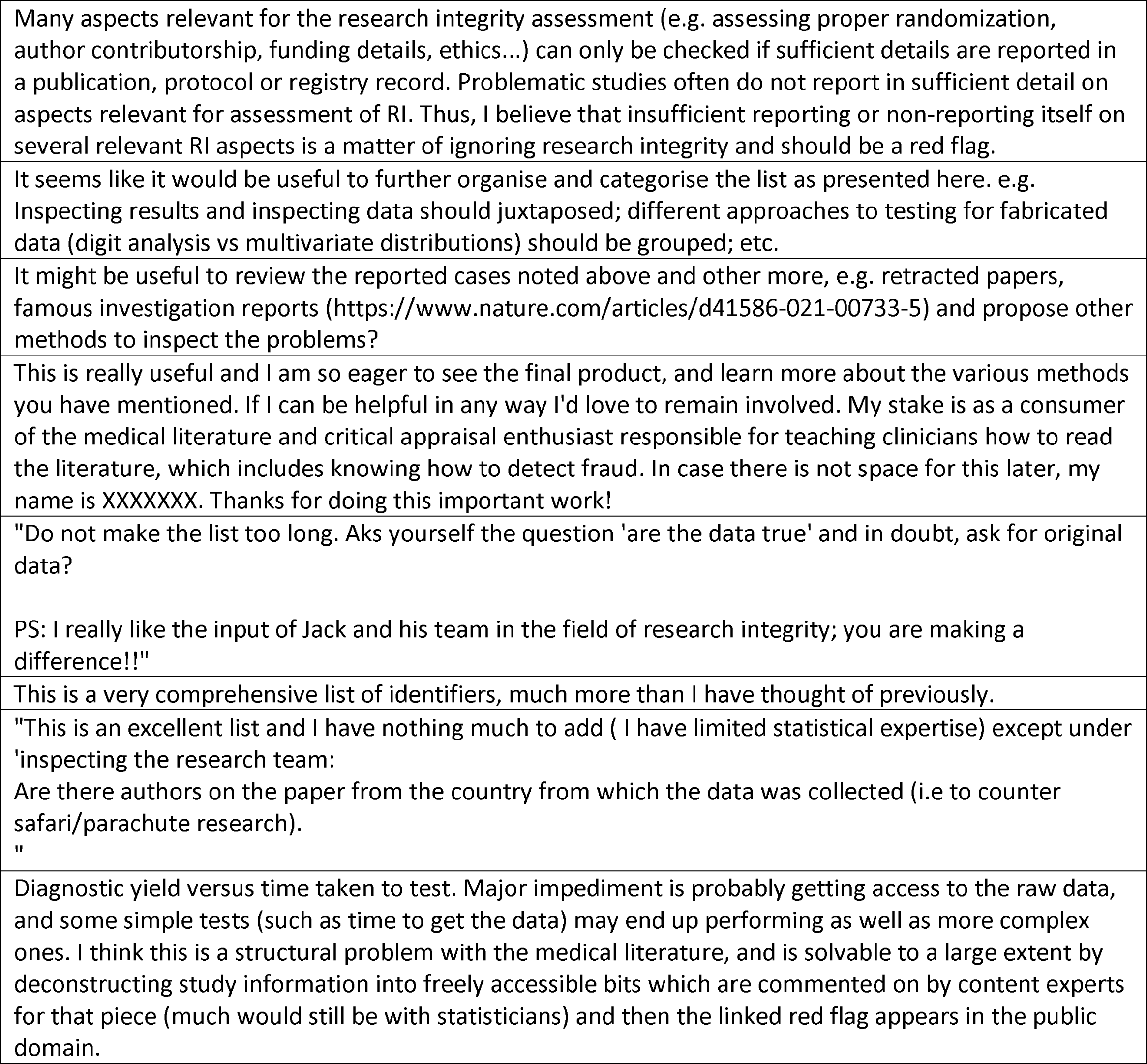

### Section B. Updated list of checks for problematic studies

Checks are arranged in five domains. Numbers in brackets/ parentheses are citation in main text

#### Domain 1: Inspecting the results in the paper (28 checks)

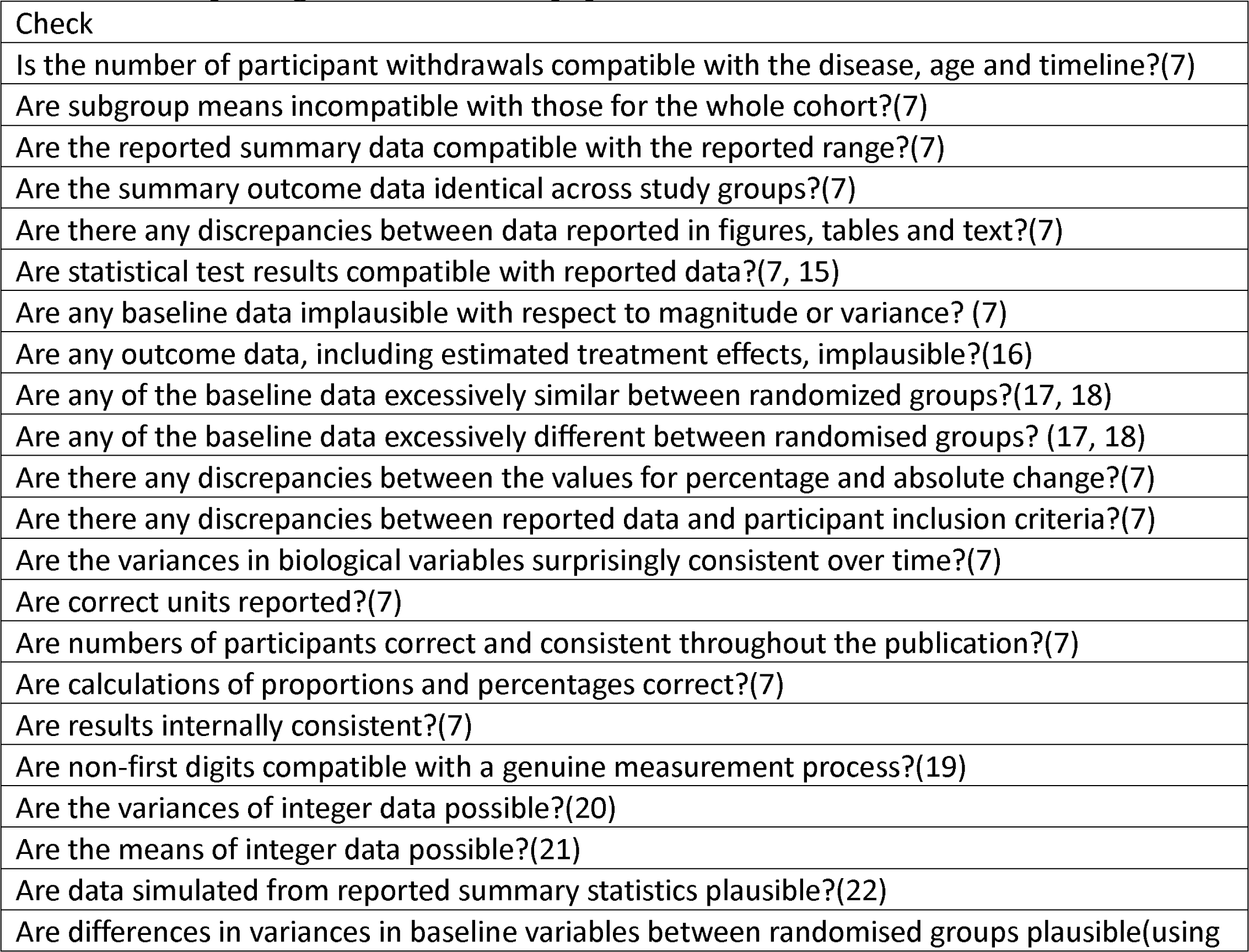
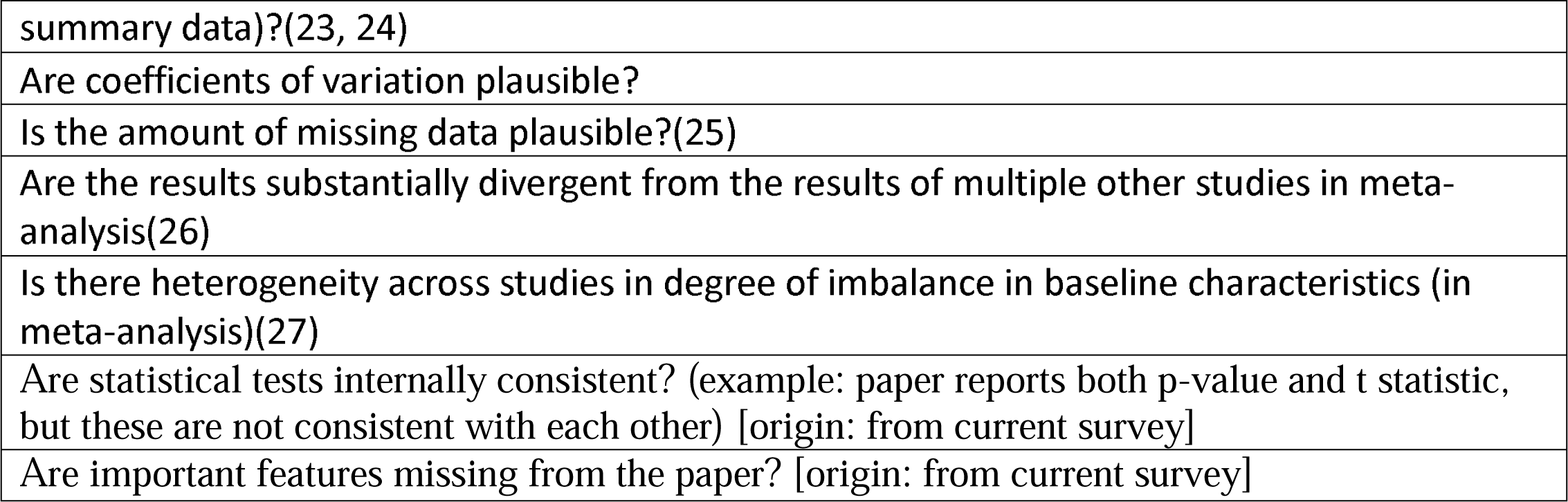

#### Domain 2: Inspecting the research team and their work (19 checks)

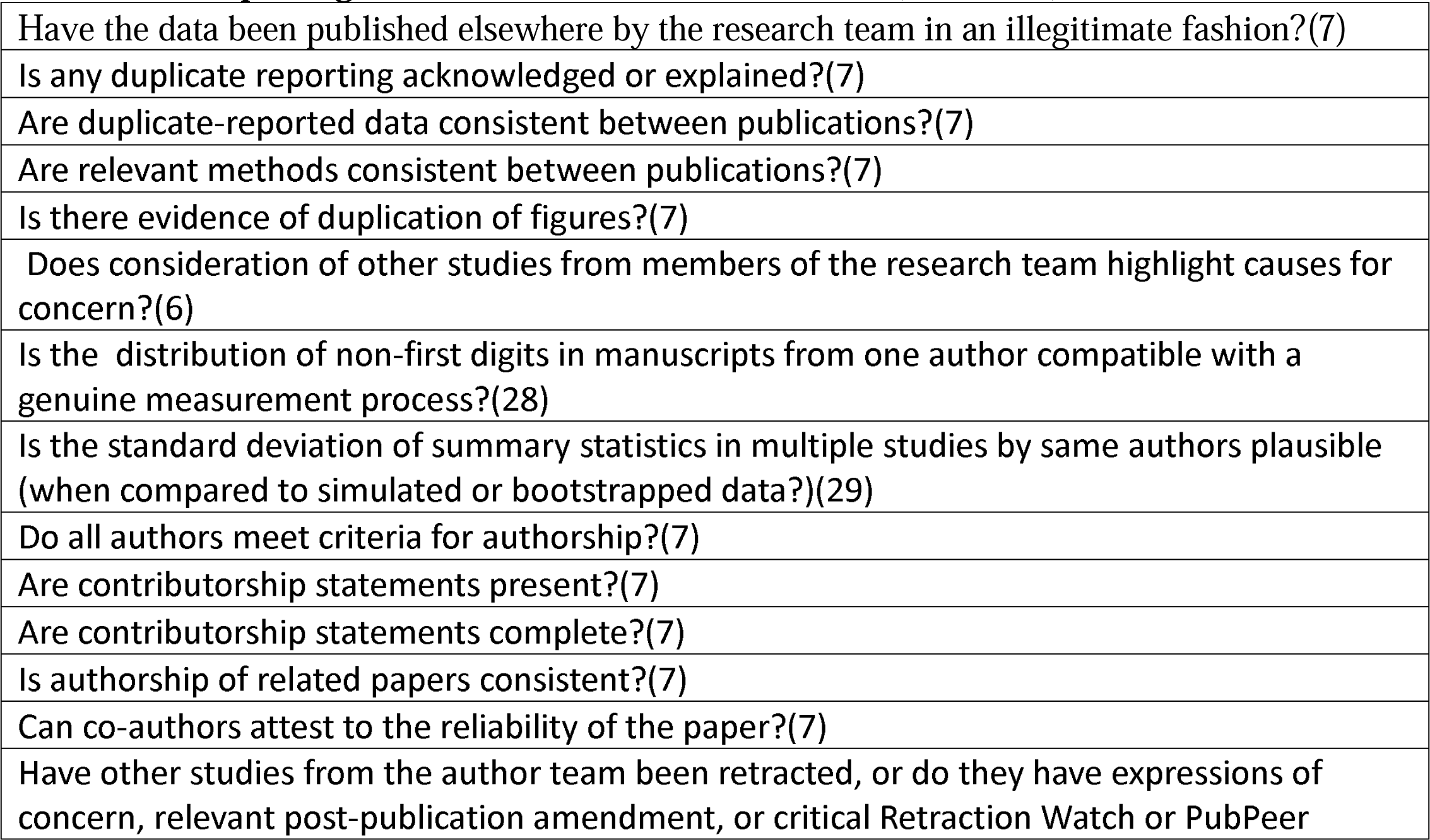
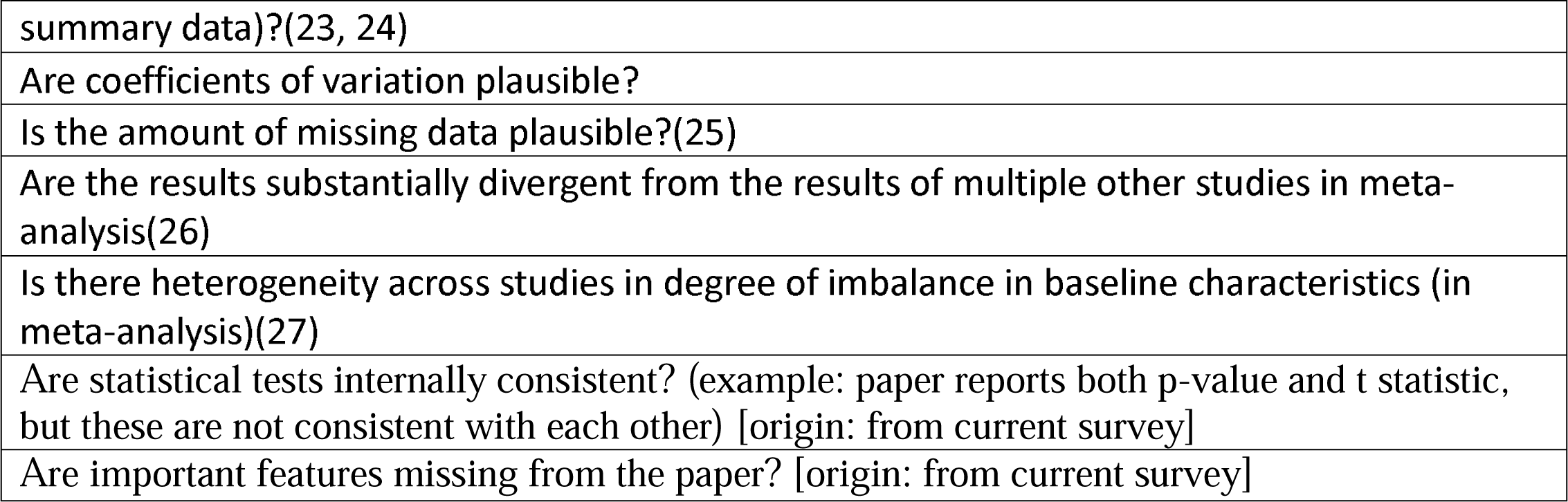

#### Domain 3: Inspecting conduct, governance and transparency (21 checks)

|  |
| --- |
| Is the volume of work reported by research group plausible, including that indicated by concurrent studies from the same group?(7) |
| Is the reported staffing adequate for the study conduct as reported?(7) |
| Is the recruitment of participants plausible within the stated time frame for the research?(7) |
| Is the recruitment of participants plausible considering the epidemiology of the disease in the area of the study location?(7) |
| Is the interval between study completion and manuscript submission plausible?(7) |
| Could the study plausibly be completed as described?(7) |
| Are the study methods plausible, at the location specified?(7) |
| Are the locations where the research took place specified, and is this information plausible?(7) |
| Is a funding source reported?(7) |
| Has the study been prospectively registered?(7) |
| Are details such as dates and study methods in the publication consistent with those in the registration documents?(7) |
| Is there evidence that the work has been approved by a specific, recognized committee? (ethics)(7) |
| Are there any concerns about unethical practice?(7) |
| Is the grant funding number identical to the number in unrelated studies?(6) |
| Are the data publically available?(6) |
| Do the authors agree to share individual participant data?(6) |
| Are additional patient data recorded in patient case records beyond what is reported in the paper?(6) |
| Do authors cooperate with requests for information? [author team suggestion] |
| Do authors provide satisfactory responses to requests? [author team suggestion] |
| In which country was the study conducted? [origin: from the current survey] |
| Is the procedure of the study aligned with local legislations? [origin: from the current survey] |

#### Domain 4: Inspecting text and publication details (8 checks)

|  |
| --- |
| Is there evidence of copied work, such as duplicated or partially duplicated tables?(7) |
| Is there evidence of text reuse (cutting and pasting text between papers), including text that is inconsistent with the study?(7, 30-34) |
| Are there typographical errors?(7) |
| Is there evidence of automatically-generated text?(35) |
| Has the study been retracted or does it have an expression of concern, a relevant post-publication amendment, a critical Retraction Watch or PubPeer comment or has been previously excluded from a systematic review?(6) |
| Was the study published in a journal from a list of predatory/ low quality journals?(6) |
| Is there evidence of manipulation or duplication of images?(7) |
| Was the time between submission to acceptance reasonable? [origin: from current survey] |

#### Domain 5: Inspecting individual participant data (40 checks)

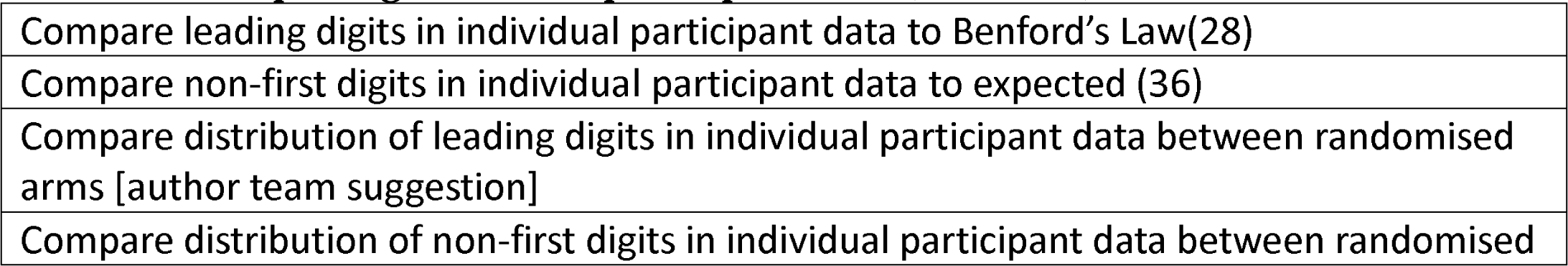
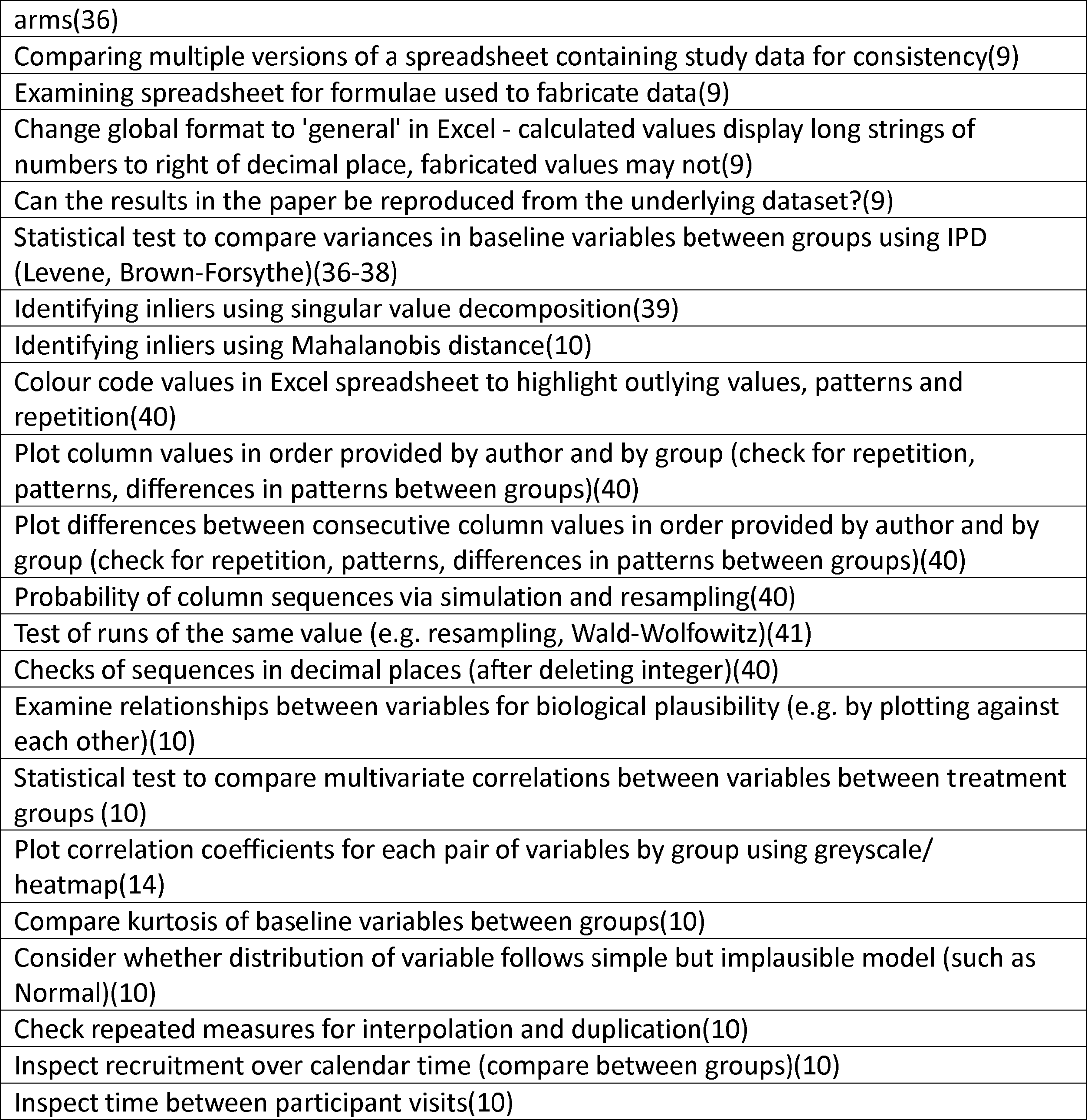
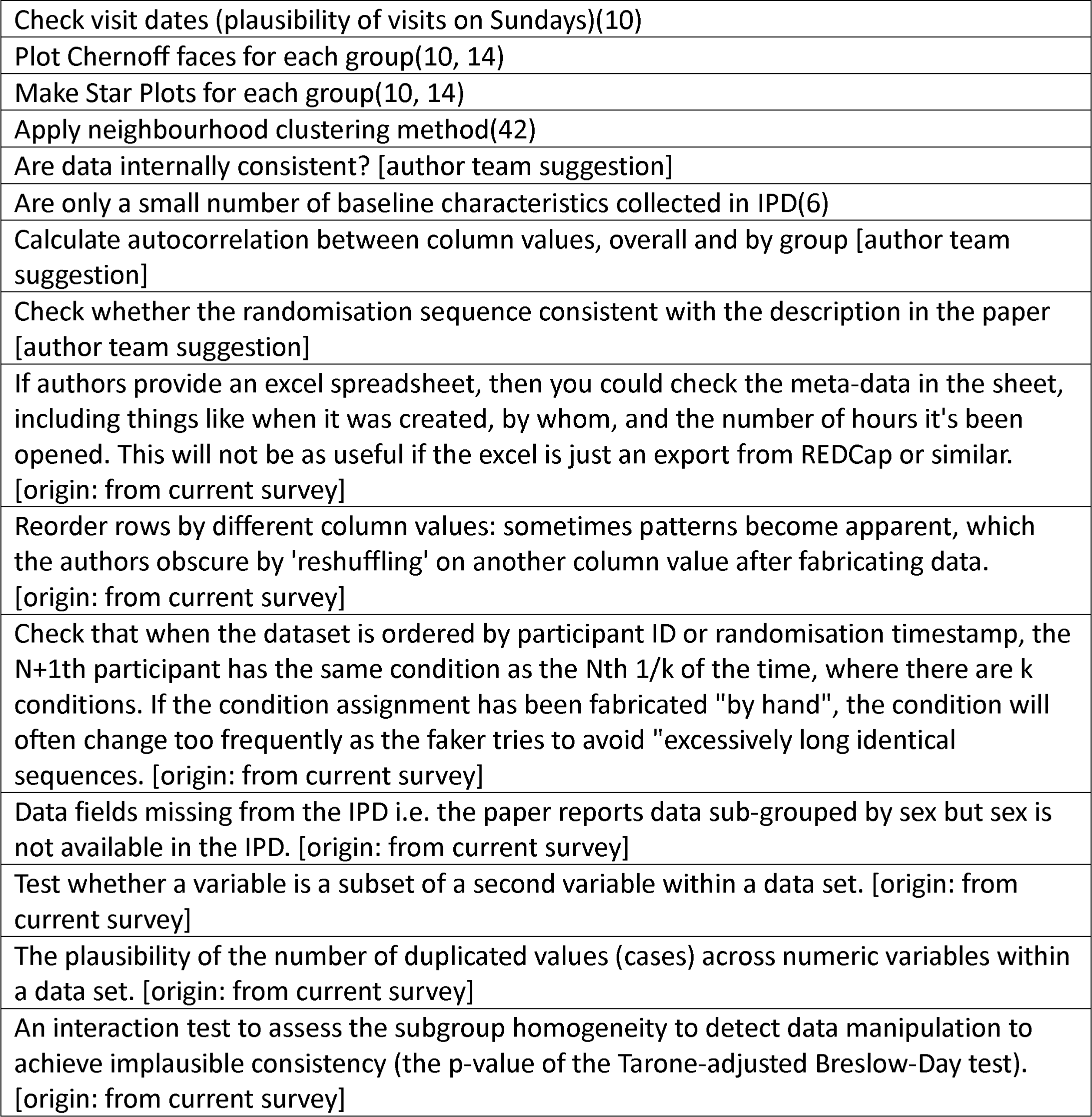

### Section C. Items excluded from the survey. Numbers I brackets/ parentheses are citation in main document

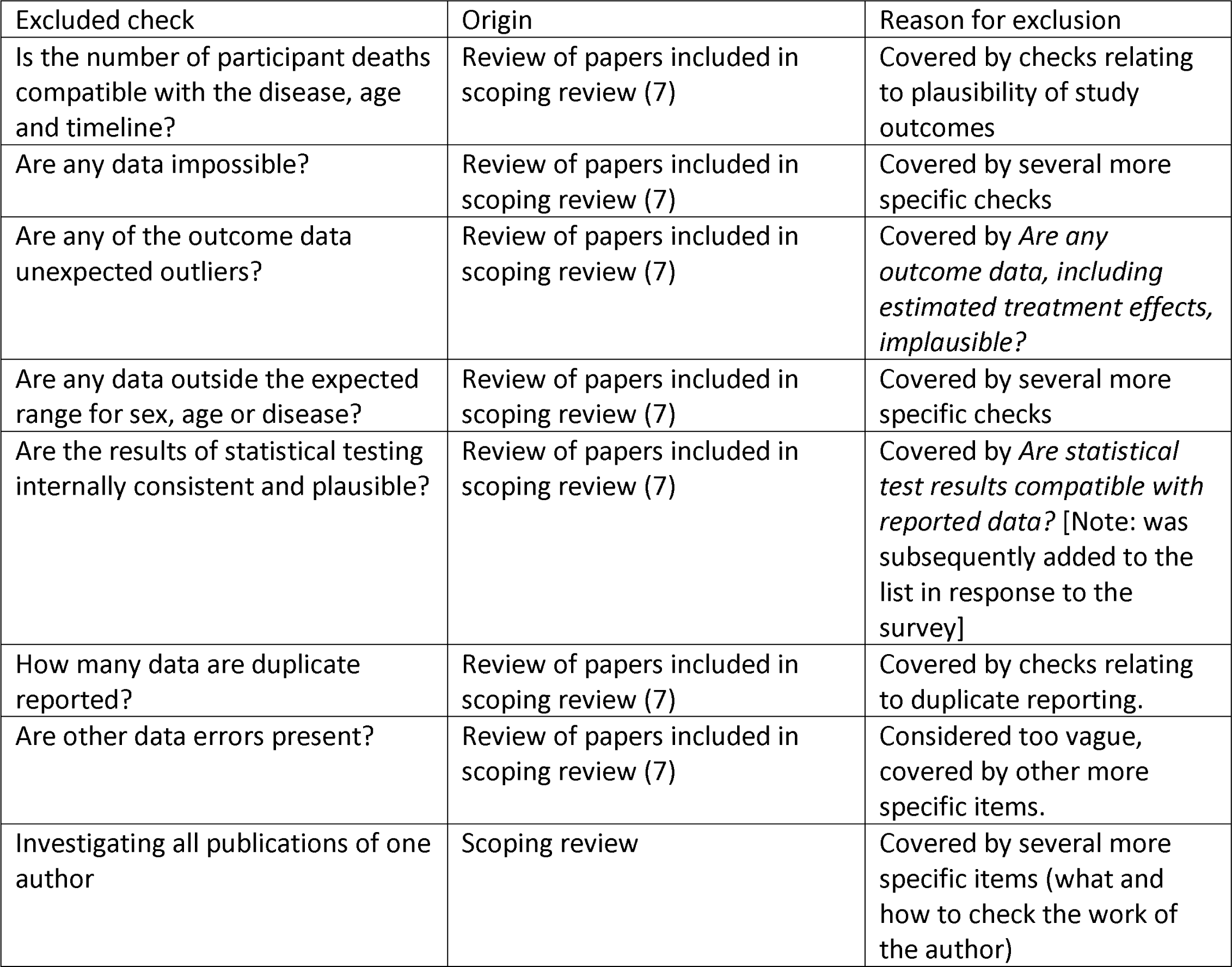
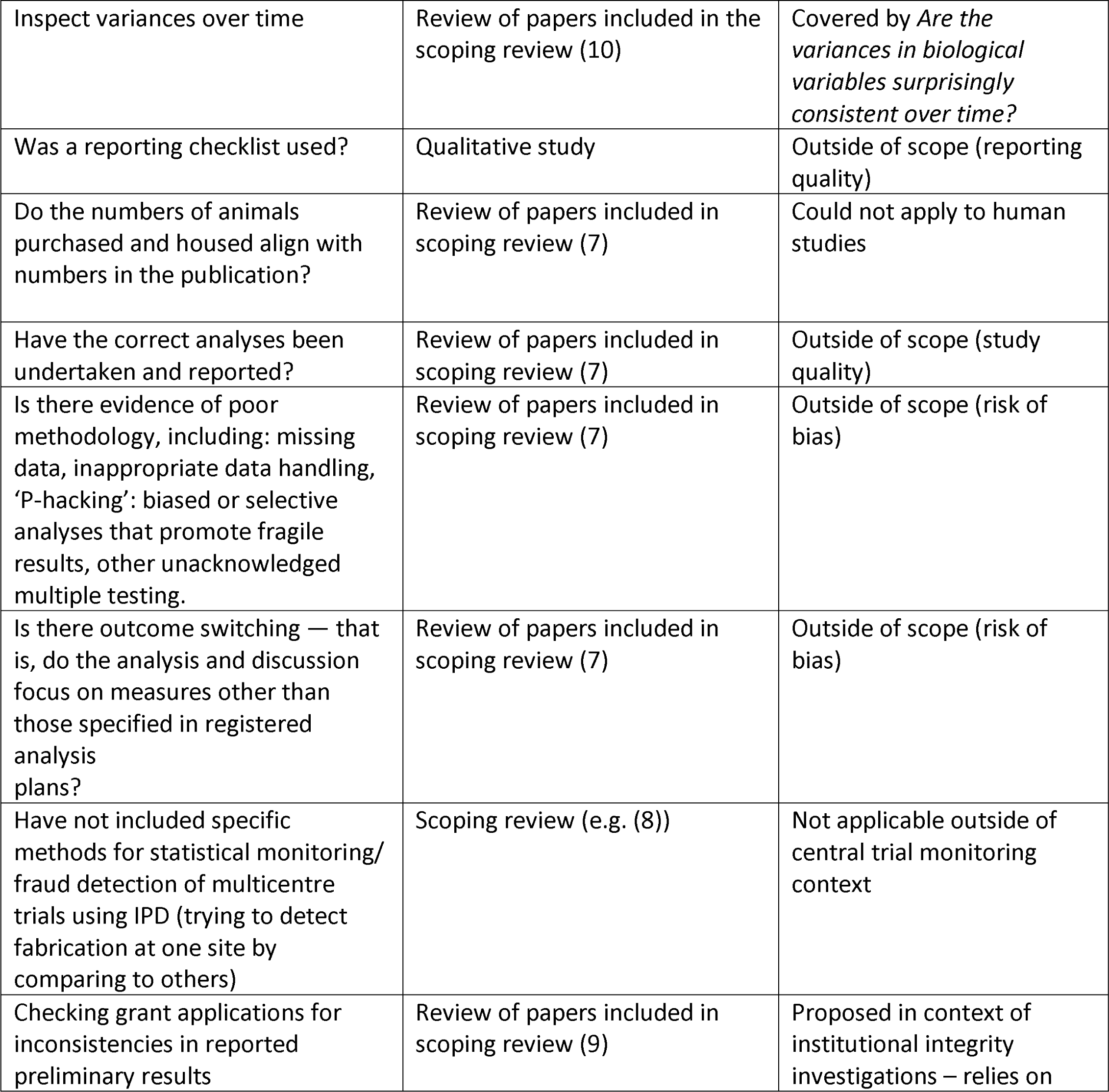
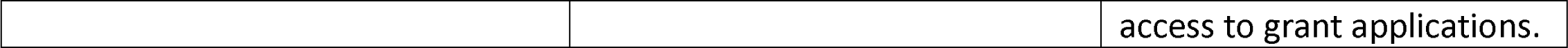

## Notes

### Clinical Protocols

https://osf.io/6pmx5/

https://bmjopen.bmj.com/content/14/3/e084164.info

### Author Declarations

The University of Manchester ethics decision tool was used on 30/09/22. Ethical approval was not required for this study, since it involved asking experts for their professional opinion. This was confirmed with the Ethics Signatory.

### Summary of Updates

Table 3 replaced with a figure (new Figure 1). Old Figure 1 has become new Figure 2. Updates to author list and declarations.

